# Genetic liability to sedentary behavior increases the risk of cardiovascular disease incidence: Evidence from the FinnGen cohort with 293,250 individuals

**DOI:** 10.1101/2024.06.20.24309213

**Authors:** L. Joensuu, K. Koivunen, N. Tynkkynen, T. Palviainen, J. Kaprio, FinnGen, M. Klevjer, K. Øvretveit, U. Wisløff, A. Bye, U. Ekelund, E. Sillanpää

**Affiliations:** Gerontology Research Center, Faculty of Sport and Health Sciences, University of Jyväskylä, Jyväskylä, FINLAND; Institute for Molecular Medicine Finland, HiLIFE, University of Helsinki, Helsinki, FINLAND; Cardiac Exercise Research Group, Department of Circulation and Medical Imaging, Faculty of Medicine and Health Sciences, Norwegian University of Science and Technology, Trondheim, NORWAY; Clinic of Cardiology, St. Olav’s Hospital, Trondheim, NORWAY; HUNT Center for Molecular and Clinical Epidemiology, Department of Public Health and Nursing, Norwegian University of Science and Technology, Trondheim, NORWAY; Norwegian School of Sport Sciences, Oslo, NORWAY; Department of Chronic Diseases, Norwegian Institute of Public Health, Oslo, NORWAY; Wellbeing Services County of Central Finland, Jyväskylä, FINLAND

## Abstract

**Background:** It is unclear how the genetics of sedentary behavior are associated with incident cardiovascular disease (CVD). We investigated the associations between genetic liability to sedentary behavior, sedentariness, and four main CVD outcomes: any CVD, hypertensive diseases, ischemic heart diseases, and cerebrovascular diseases.

**Methods:** Leisure screen time was used as a proxy for sedentary behavior. We developed a polygenic score for leisure screen time (PGS LST) based on over 890,000 genetic variants. We tested the validity of this score against self-reported LST in the older Finnish Twin Cohort (FTC, N=2,689, mean age of 60.5±3.7 years, 54.7% women) using linear regression. We examined the associations between PGS LST and register-based records of CVDs using survival models among FinnGen participants (N=293,250–333,012, 67.0±13.0 years at follow-up, 52.3% women). We replicated analyses in an independent cohort (Trøndelag Health Study [HUNT], N=35,289, 64.0±13.1 years, 51.6% women) and explored if the associations persist following adjustments for socioeconomic status, body mass index, and smoking or are mediated via reduced physical activity.

**Results:** In the FTC, each standard deviation increase in PGS LST was associated with greater self-reported LST (hours/day) (β = 0.09, 95% CI: 0.05–0.14). In FinnGen, each standard deviation increase in PGS LST was associated with a higher risk of incident CVD (hazard ratio: 1.05, [1.05–1.06]) (168,770 cases over 17,101,133 person-years).The magnitudes of association for three most common CVDs were 1.09 (1.08–1.09), 1.06 (1.05–1.07), and 1.05 (1.04–1.06) for hypertensive diseases, ischemic heart diseases, and cerebrovascular diseases, respectively. Those in the top decile of PGS LST had 21%, 35%, 26%, and 19% higher risk of any CVD, hypertensive diseases, ischemic heart diseases, and cerebrovascular diseases, respectively, than those in the bottom decile. Associations replicated in HUNT and remained independent of covariates except for cerebrovascular diseases. Besides direct effects, reduced physical activity served as a potential mediating pathway for the associations.

**Conclusions:** A higher genetic liability to sedentary behavior is associated with a greater risk of developing CVDs, although effect sizes with current PGS remain small. Our findings suggest that genetic liability to sedentary behavior is an underrecognized driver of common CVDs.

**Clinical perspective:** What is new?

- It is not known whether a genetic liability to sedentary behavior is a mutual underlying factor for both sedentary behavior and incident cardiovascular disease at the population level.
- We observed that a higher polygenic score for leisure screen time was associated with more self-reported leisure screen time and a higher risk of common cardiovascular diseases.

What are the clinical implications?

- This study provides novel insights into the relationship between genetic predisposition to sedentary behavior and the development of cardiovascular diseases, shedding light on a previously underexplored aspect of disease etiology.
- These results may motivate health professionals to encourage sedentary persons to undertake at least some physical activity.

## Introduction

Cardiovascular diseases (CVDs) are the leading cause of disease burden and death worldwide.^1^ Sedentary behavior is any waking behavior in a sitting, reclining, or lying posture and characterized by an energy expenditure of less than or equal to 1.5 metabolic equivalents.^2^ Adults spend up to 60% of their awake time sedentary.^3^ The prevalence of sedentary behaviors has remained stable or even increased during the last 20 years.^4^

Sedentary behaviors are systematically associated with increased risk of CVD incidence and related mortality in prospective observational cohort studies.^5,6^

Interestingly, energy-saving sedentary behavior has been suggested to be a trait selected by evolution to ensure reproduction and survival.^7^ Thus, genetics may be an important driver of sedentary behavior, as supported by previous evidence from twin and candidate gene studies.^8^ However, the potential effects of this genetic liability on morbidity are unclear. From an evolutionary perspective, it is plausible that genetic predisposition to sedentary behavior increases the risk of common non-communicable diseases.^9,10^ It has been suggested that the rapid environmental changes in societies, particularly in Western countries, may have led to an “evolutionary mismatch”.^9^ In this scenario, the previously advantageous alleles, e.g. the genetic liability for sedentary behavior, may cause morbidity in the current post-industrial environment.^11^ Furthermore, as life expectancy has increased, the deleterious effects of underlying genetics may become more apparent.^10^ The theory of antagonistic pleiotropy suggests that some genes that have favored survival during reproductive ages in the past may currently limit lifespan and increase morbidity.^12^ Underlying genetics may therefore be an important confounding factor in the commonly observed associations between sedentary behavior and incident CVD.

Polygenic scores (PGSs) summarize the genome-wide information between single-nucleotide polymorphisms (SNPs) and a phenotype into a single variable that quantifies a person’s genetic liability for a given trait.^13^ In this study, we assess whether a higher genetic liability for sedentary behavior is associated with an increased risk of incident CVD at the population level. Leisure screen time (LST) is a commonly reported voluntary mode of sedentary behavior and thus serves as a proxy for this trait.^6^ We hypothesized that higher genetic liability for LST, as measured by a polygenic score (PGS LST), is associated with an increased risk of developing a CVD.

## Methods

This study consists of four phases. First, we constructed a PGS for LST. Second, we assessed the validity of this score against self-reported LST in the older Finnish Twin Cohort (FTC). Third, we assessed the associations between the PGS LST and incident CVD in the FinnGen cohort (∼10% of the Finnish population). Finally, we replicated the analyses in an independent cohort (The Trøndelag Health Study [the HUNT study]). The study design and cohorts are shown in Figure 1. Additionally, we conducted exploratory analyses to estimate the potential explanations for the observed associations.

**Figure 1.**
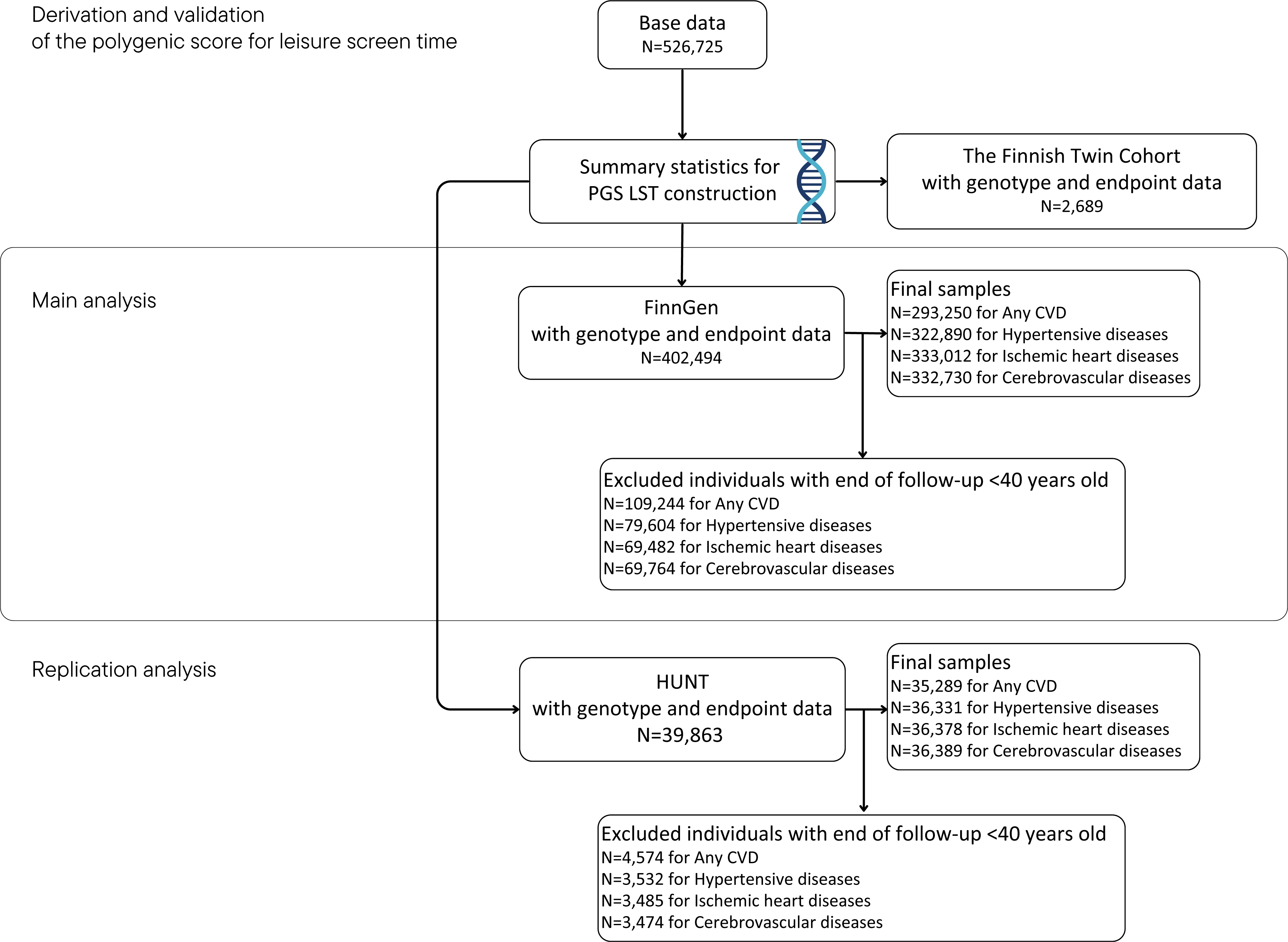
Study design and cohorts. PGS LST, polygenic score for leisure screentime; CVD, cardiovascular disease.

### Study cohorts

#### Base data for PGS LST

We used the publicly available genome-wide association study (GWAS) summary statistics previously published by Wang et al. (2022) as the base data for PGS LST calculation. The data included 526,725 genotyped individuals from 24 different cohorts with European ancestry (mean age of 49.5 ± 7.6 years). For more information regarding the base data, please see Table S1.

#### Validation cohort

The FTC is a prospective population-based cohort study that began in 1975 and has provided a 50-year follow-up resource for genetic epidemiology. The study details have been described elsewhere, but briefly, all same-sex twins born before 1958 and residing in Finland were invited to participate in the study.^14^ The majority of the invitees accepted (response rate of 84.4%, N = 31,145). In this study, we used a genotyped sub-sample of the FTC who completed a questionnaire on their sedentary behavior in 2011 (N = 2,689, mean age of 60.5 ± 3.7 years, 54.7% women) (Table S2).

#### Main cohort

FinnGen is a public–private partnership research project linking genotype and digital health record data from Finnish health registries using unique national personal identification numbers. The FinnGen data release 11 comprises 520,210 biobank participants (https://www.finngen.fi/en).^15^ The biobanks include regional, hospital-based biobanks (Auria Biobank, Biobank of Central Finland, Biobank of Eastern Finland, Borealis Biobank, Helsinki Biobank, Tampere Biobank), the Blood Service Biobank, the Terveystalo Biobank, and the Finnish Institute for Health and Welfare (THL) Biobank, which includes phenotypic and genetic information from the following studies: Botnia, Corogene, FinHealth 2017, FinIPF, FINRISK 1992–2012, GeneRisk, Health 2000, Health 2011, Kuusamo, Migraine, Super, T1D, and Finnish Twin Cohort. In our study, we excluded participants under the age of 40 years at the end of the follow-up. This was done to focus on a more homogenous adult population and CVDs, which develop over time. This process resulted in a sample of 293,250 participants for incidence of any CVD. We used the same criteria to obtain samples for the three most prevalent CVDs in the FinnGen, with 322,890 participants for hypertensive diseases, 333,012 for ischemic heart diseases, and 332,730 for cerebrovascular diseases. The larger sample sizes for the three most common CVDs result from their later onset in life compared to all potential CVD diagnoses.

#### Replication and exploratory cohort

The HUNT study is conducted in the Trøndelag region of Norway. It commenced in 1984, inviting all residents 20 years old and above in the former county of Nord-Trøndelag, now Trøndelag, to participate. To date, a total of four surveys have been completed: HUNT1 (1984–1986, N = 77,202, 89.4% participation rate), HUNT2 (1995–1997, N = 65,228, 69.5% participation rate), HUNT3 (2006–2008, N = 50,800, 54.1% participation rate), and HUNT4 (2017–2019, N = 56,04, 54.0% participation rate).^16^ The study data include laboratory measurements, questionnaires, interviews, and clinical examinations performed by trained personnel. More information about HUNT is available on the project’s website (*<u>ntnu.edu/hunt</u>*). In this study, we used a subsample of individuals from HUNT2 and HUNT3 with available genotype and health record data (N = 35,289).

### Ethical considerations

The base data for PGS LST were published in the National Human Genome Research Institute-European Bioinformatics Institute GWAS Catalog. The catalog follows the General Data Protection Regulation applicable from 25 May 2018.^17^ The different processes of the FTC study and data pooling have been evaluated by ethics committees of the University of Helsinki (113/E3/01 and 346/E0/05), Helsinki University Central Hospital (136/E3/01, 01/2011, 270/13/03/01/2008, 154/13/03/00/2011, HUS/1799/2017), and the Finnish Institute for Health and Welfare (THL/4743/6.02.04/2021). The FinnGen cohort consists of participants from Finnish biobanks. Participants in FinnGen provided informed consent for biobank research on the basis of the Finnish Biobank Act. Alternatively, separate research cohorts, collected before the Finnish Biobank Act came into effect (September 2013) and the start of FinnGen (August 2017), were collected based on study-specific consent and later transferred to the Finnish biobanks after approval by Fimea, the National Supervisory Authority for Welfare and Health. Further information related to ethical procedures and permits in the FinnGen study are provided in the Supplemental Material (under Supplemental Methods, page 2). The HUNT study was approved by the Regional Committee for Medical and Health Research Ethics (REC; 2019/29771), the Trøndelag Health Study, the Norwegian Data Inspectorate, and the National Directorate of Health. All participants provided written informed consent. The authors assume responsibility for the accuracy and completeness of the protocol, data, and analyses, fidelity of their reporting, and compliance with the Declaration of Helsinki, national laws, and guidelines of the Finnish Advisory Board on Research Integrity.

### Outcomes

#### LST

For the validation analyses in the FTC, LST in hours per day (h/day) was used as the outcome and included self-reported television and video viewing and computer use at home. This outcome was selected to match exactly the outcome used in the base data for PGS LST. In the FTC, participants reported how many hours per day they spent sitting: at home watching TV or videos and at home at the computer. Each question had four choices: (a) less than an hour, (b) an hour–less than two hours, (c) two hours–less than four hours, and (d) four hours or more.^18^ Answers were summarized to indicate LST (h/day). The data were collected in 2011 and represent screen time activities at that time.

#### CVD

The CVD categorization was based on FinnGen endpoint definitions. The following ICD-10 codes (ICD-9 codes [and FinnGen labels]) were used: All CVDs, I00-I99, T82.2, Z95.1 (390-459, 4019X, 4029A, 4029B, 4039A, 4040A, 4059A, 4059B, 4059X, 4160A, 4330A, 4330X, 4331A, 4331X, 4339A, 4339X, 4340A, 4341A, 4349A, 4371, 4372, 4372A, 4373, 4374, 4375, 4376, 4378X, and 9960A [I9_CVD]); Hypertensive diseases, I10-I15, I27.0, I67.4 (4019X, 4029A, 4029B, 4039A, 4040A, 4059A, 4059B, 4059X, 4160A, and 4372A [I9_HYPERTENSION]), Ischemic heart diseases, I20-I25, T82.2, Z95.1 (410, 411[0-1], 412, 413, 414, 9960A [I9_ISCHHEART]), and Cerebrovascular diseases, I60-I69 (430, 431, 436, 438, 4330A, 4330X, 4331A, 4331X, 4339A, 4339X, 4340A, 4341A, 4349A, 4371, 4372, 4373, 4374, 4375, 4376, and 4378X [I9_CEREBVASC]). The register data were based on the national hospital discharge (available from 1968) and causes of death (available from 1969) registers. Additionally, any CVDs included data from Care Register for Health Care for coronary angioplasty, coronary artery bypass, valvular operations, endovascular or surgical operations to intracerebral aneurysms, and peripheral artery operations and from the Finnish Social Insurance Institution drug reimbursement register for atrial fibrillation and flutter, heart failure, and hypertension. Hypertensive diseases also included data from the drug reimbursement register. Further descriptive information, such as disease prevalence and age distribution, can be queried using FinnGen labels via the Risteys interface (<u>risteys.finregistry.fi</u>). We utilized the same ICD-10 and ICD-9 codes in the HUNT replication analyses. The HUNT health records are based on the Nord-Trøndelag Health Trust discharge register (available from 1987).

### Exposure

The PGS LST was based on the largest available GWAS meta-analysis for LST at the time of the analysis (GWAS Catalog Study ID: GCST90104339).^19^ In this data, the LST included self-reported television viewing, video game playing, and computer use at home. The discovery data reportedly explained 2.8% of the variance in self-reported LST,^19^ and the SNP heritability was 7.4%.^20^ The base data did not overlap with the later used FTC, FinnGen or HUNT study cohorts.

We used the high-performing SbayesR approach to construct the PGS.^21^ In SbayesR, the base data summary statistics are re-weighted to consider the linkage disequilibrium between each variant and restricted to HapMap3 SNPs to ensure computational efficiency while representing the whole genome. The pipeline and relevant codes are available elsewhere.^22^ The final number of processed variants was 890,575 in the FTC, 891,628 in the FinnGen cohort, and 901,640 in the HUNT cohort. We used standardized PGS LST scores (mean of zero, standard deviation of one) in the analyses. The PGS LST fitted well to the FTC, FinnGen, and HUNT cohorts and followed a normal distribution.

### Genotyping, quality control, and imputation

In the FTC, chip genotyping was done using Illumina Human610-Quad v1.0 B, Human670-QuadCustom v1.0 A, Illumina HumanCoreExome, and Affymetrix FinnGen Axiom arrays. Genotype quality control was done in three batches (batch1: 610k+670k chip, batch2: HumanCoreExome, and batch3: Affymetrix chip genotypes), and genotypes of all batches were imputed to the Haplotype Reference Consortium release 1.1 reference panel.^23^ More details related to genotyping procedures in the FTC are available in the Supplemental Material (Supplemental Methods, page 2).

The FinnGen samples were genotyped with Illumina and Affymetrix chip arrays (Illumina Inc., San Diego, and Thermo Fisher Scientific, Santa Clara, CA, USA). A detailed information on genotyping, quality control, and imputation has been described elsewhere: https://finngen.gitbook.io/documentation/.

In the HUNT study, a total of ∼69,000 participants contributed with DNA for genotyping in HUNT2 and HUNT3. Detailed information on genotyping, imputation, and quality control is described in more detail elsewhere.^24^

### Covariates

Covariates were based on previous literature.^25,26^ The first ten principal components of ancestry adjust for any genetic stratification that may occur in the study population and were used alongside with sex as a covariate in all analyses.^25^ Additional covariates were specific to the analysis. In validation analyses in the FTC, we also used age, educational attainment (highest achieved self-reported educational degree in the Finnish educational system; low, basic education degree at most; middle, basic education and additional studies; and high, at least upper secondary degree),^27^ and body mass index (BMI) based on valid self-reported height and weight.^28^ In main and replication analyses, we also used genotyping batch, which accounts for the potential stratification that may occur form the genotyping array and sample. In further exploratory analyses with the replication cohort, we controlled for variance caused by highly plausible covariates, such as BMI (based on measured height and weight [Jenix DS-102, Dong Sahn Jenix Co, Ltd, Korea]), socioeconomic status (SES; derived from occupational status),^26^ and self-reported smoking status (never, former, current) using the HUNT3 data.

### Statistical analysis

#### Validation analyses

All validation analyses were performed with Stata (StataCorp, College Station, TX, USA). Correlation between PGS LST and self-reported LST was tested with Pearson correlation. The association between PGS LST and self-reported LST was assessed with linear regression in the FTC, where the twin structure of the data was acknowledged using the svyset command (i.e., the twin pair was the primary sampling unit). The crude model included standardized PGS LST and the first ten principal components of ancestry. Model 1 was adjusted additionally for age and sex, and Model 2 was further adjusted with educational attainment and BMI. Differences between high and low genetic liabilities were evaluated using linear regression, where a binary variable indicated whether the individual belonged to the highest or lowest decile of PGS LST. A priori power calculations indicated that sufficient statistical power would be reached with a sample size of N > 950 (expected small effect size 0.02, 15 predictors, power at 0.8 level, and α set to 0.05).

#### Main analyses

We used Cox proportional hazard models and R packages survival^29^ and survminer^30^ to estimate the hazard ratios (HRs) and 95% confidence intervals (CI) between PGS LST and incident CVD in the FinnGen cohort. Visual inspection of Schoenfeld residuals and log-minus-log plots indicated that the finally adjusted models satisfy the proportional hazard assumptions.^31^ PGS×SEX interaction was tested, but no replicable sex differences were observed; thus, analyses were conducted for men and women in combination. We used age as the time scale and the first ten principal components of ancestry, genotyping batch, and sex as covariates. Start of follow-up was set at birth, as the PGS remains stable from conception. Follow-up ended with whichever came first among the first record of the endpoint of interest, death, or the end of follow-up on 31 December 2021. Utilizing the weights of the European standard population, age-standardized incidence was calculated when relevant.^32^ We also compared the hazard ratios (relative measures of survival) and cohort-specific cumulative incidence of events (absolute measures of survival) between the highest and lowest deciles of PGS LST. We additionally conducted competing risk analysis with Fine-Gray sub-distribution hazard models for the three main CVDs, as one event may preclude the occurrence of another. Here, we coded censored as “0,” events of interest as “1,” and other competing CVD events as “2”.^33^ Additionally, as the exposure (PGS LST) might affect who survives until genotyping and also to control for immortal time bias, we conducted sensitivity analyses in which the start of the follow-up was set to the date of individual DNA sampling. Sufficient statistical power was expected to be achieved with > 120 cases, using a classical definition of 10 cases per predictor variable.

#### Replication analyses

The replication analyses were conducted in HUNT and followed a similar procedure as the main survival analyses, except that when mortality data were not available, the end of follow-up was assigned to whichever came first between the first record of the endpoint of interest and last verified contact with healthcare.

#### Exploratory analyses

In HUNT, we also conducted several exploratory analyses to assess the potential sources of observations in the main and replication analyses. We tested if the associations were independent of highly plausible covariates with a complete case subsample of HUNT3. In this analysis, Model 1 included PGS LST, the first ten principal components of ancestry, sex, and genotyping batch while Model 2 included additionally SES, Model 3 additionally BMI, and Model 4 additionally smoking status. We also tested a potential mediating pathway from PGS LST to incident CVD via sedentary behavior using the product of coefficients approach and accompanying Sobel test, which are suitable for survival analysis with common outcomes.^34^ In this mediation analysis, we used participants from the sensitivity analyses with self-reported total sitting time (h/day): “About how many hours do you sit during an average day? (include work hours and leisure time)”. Only diseases for which a direct association between PGS LST and incident disease was observed in the sensitivity analyses were included in the mediation analysis. Association between PGS LST and total sitting time were evaluated with linear regression, while a Cox proportional hazards model was used for the association between total sitting time and disease incidence. The models were adjusted for sex similar to prior sensitivity analyses but not for the first ten principal components or genotyping batch for consistency across pathways. These mediating analyses were replicated with a physical activity variable, i.e., metabolic equivalent (MET) hours per week, calculated as the product of self-reported physical activity frequency, intensity, and duration. Across all analyses, all tests were two-tailed, and α was set at p < 0.05.

## Results

### Association between polygenic score for leisure screen time and self-reported leisure screen time

The 2,689 individuals of the FTC reported an average of 3.9 ± 1.1 hours LST per day (Table S2). The effect size for PGS LST against LST was small (Pearson correlation coefficient = 0.08, p < 0.001), and PGS LST explained 0.7% of the variance in the LST. PGS LST of one standard deviation higher was associated with higher self-reported LST (h/day), with β = 0.09 (95% CI: 0.05–0.14) in crude models; β = 0.09 (95% CI: 0.05–0.13) after adjustments for age, and sex; and β = 0.08 (95% CI: 0.04–0.13) after further adjustments for educational attainment and BMI (Table S3). Correspondingly, individuals in the highest decile for PGS LST reported an average of 4.1 ± 1.0 h/day LST, while those in the lowest decile reported an average of 3.7 (1.1) h/day (p < 0.001 for difference) (Table S4).

### Association between polygenic score for leisure screen time and cardiovascular disease incidence

Descriptive data on 293,250 participants in the FinnGen sample of any CVD are shown in Table 1. Fifty-eight percent in the FinnGen cohort were diagnosed with a CVD in adulthood (≥ 40 years old, 168,770 cases) with an age-standardized incidence of 34.9 per 10,000 person-years (Figure 2). The average follow-up time from birth was 58 (40 to 106) years, with a total of 17,101,133 person-years. The descriptives of 35,289 participants in the HUNT replication cohort are presented in Table 1. The age-standardized incidence for any CVD was 36.5 per 10,000 person-years (Figure 2), with an average follow-up time of 64 (40 to 103) years and a total of 2,257,493 person-years.

**Figure 2.**
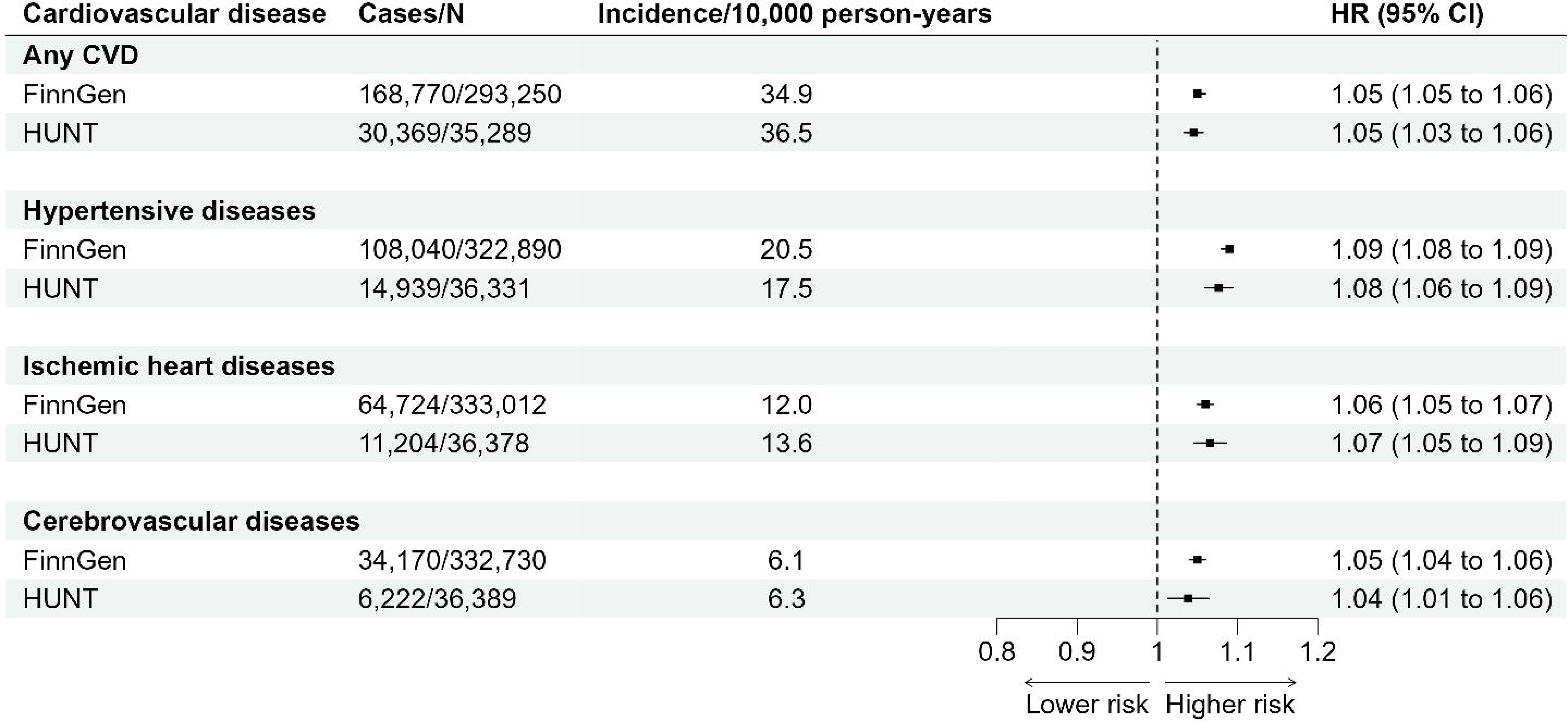
Associations between polygenic score for leisure screen time and cardiovascular disease incidence. Values are hazard ratios (HRs) and 95% confidence intervals (CIs) per one standard deviation increase in the polygenic score for leisure screen time. We used age as the time scale, and the first ten principal components of ancestry, sex, and genotyping batch as covariates; CVD, cardiovascular disease. Incidence values are age-standardized according to the European Standard Population to enable comparison between FinnGen and HUNT cohorts.

**Table 1.** Descriptives of individuals with any CVD in the main FinnGen cohort and in the replication cohort (HUNT).

| Characteristic | Main cohort |  | Replication cohort |  |
| --- | --- | --- | --- | --- |
|  | N | FinnGen | N | HUNT |
| Age (years) | 293,250 | 67.0 (13.0) | 35,289 | 64.0 (13.1) |
| Female (%) | 293,250 | 52.3 | 18,218 | 51.6 |
| BMI (kg/m<sup>2</sup>) | 82,224 | 27.8 (5.3) | 22,398 | 27.9 (4.5) |
| Smoking status | 48,391 |  | 21,768 |  |
| Never (%) | 17,915 | 37.0 | 7,921 | 36.4 |
| Former (%) | 7,248 | 15.0 | 8,607 | 39.5 |
| Current (%) | 23,228 | 48.0 | 5,240 | 24.1 |

Values are presented as means (standard deviation) for continuous variables and percentages for others. Age is from the end of follow-up, and others the last known information during adulthood; BMI, body mass index; FinnGen data is from FinnGen data release 11.

In FinnGen, PGS LST of one standard deviation higher was associated with a higher risk of any CVD after adjusting for the first ten principal components of ancestry, sex, and genotyping batch (HR: 1.05, 95% CI: 1.05–1.06; Figure 2). Disease-specific associations for three most common CVDs were 1) 1.09 (1.08 – 1.09) for hypertensive diseases (108,040 cases), 2) 1.06 (1.05 – 1.07) for ischemic heart diseases (64,724 cases), and 3) 1.05 (1.04– 1.06) for cerebrovascular diseases (34,170 cases). The associations were replicated in the HUNT cohort with exactly or very similar effect sizes (Figure 2).

In the FinnGen cohort, those in the highest PGS LST decile had 19% to 35% higher risk of CVDs compared to those in the lowest PGS LST decile (Table S5). For example, at 60 years of age, the cumulative incidence for any CVDs was 44% for those with high PGS LST, while for those with low PGS LST it was 37% (Table S7, Figure 3). This higher risk of incident CVD in participants with higher genetic liability replicated in HUNT, except for cerebrovascular disease (HR: 1.08 [0.96–1.20]) (Table S6, Figure S1).

**Figure 3.**
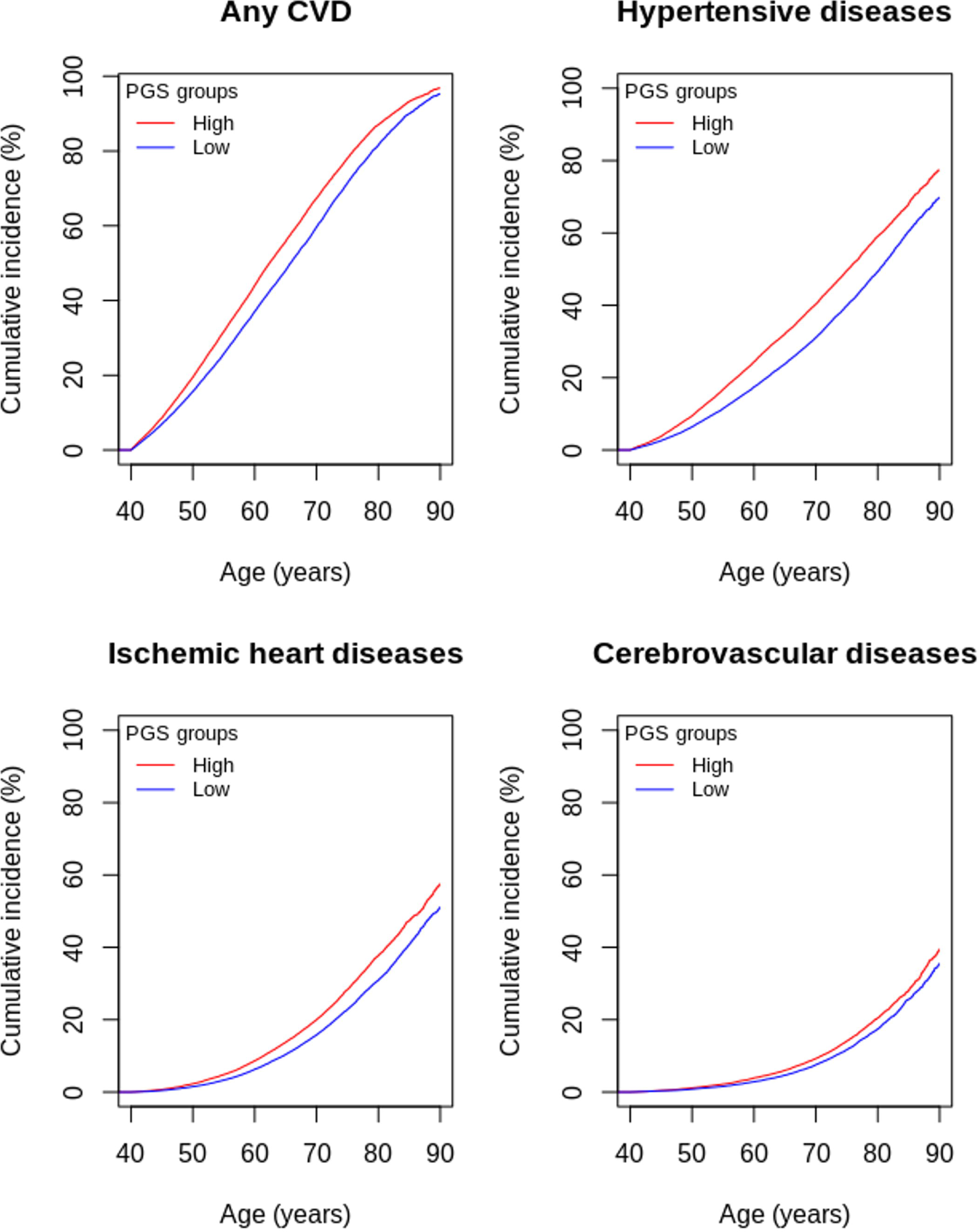
Cumulative incidence curves for high (> 90^th^ percentile [red]) and low (< 10^th^ percentile [blue]) PGS LST groups in the FinnGen cohort based on Cox proportional hazard models and adjusted for the first ten principal components, sex, and genotyping batch.

Considering other CVDs as competing events attenuated the cumulative incidence estimates for both high and low PGS LST participants, but their difference remained statistically significant for hypertensive diseases and ischemic heart diseases, although not for cerebrovascular diseases (Table S8). In further sensitivity analyses with follow-up starting from individual DNA sampling dates (mean follow-up was 6.0 years in FinnGen and 5.0 in HUNT), the associations remained the same or were slightly attenuated in the FinnGen cohort, and replicated in the HUNT cohort except for any CVD and cerebrovascular diseases (Table S9).

Exploratory analyses showed that the association between PGS LST and incident CVD persisted after adjustments for SES, BMI, and smoking status, except for cerebrovascular diseases, where the number of cases was smallest (Figure S2). Mediation analysis showed that higher PGS LST was associated with higher self-reported total sitting time (h/day) in the HUNT cohort (β = 0.06, standard error [SE] = 0.02, p = 0.016 in a model for hypertensive diseases and β = 0.07, SE = 0.02, p = 0.003 for ischemic heart diseases) but indicated no potential mediating pathway to incident CVD (Figure S3). However, higher PGS LST was also associated with lower MET h/week (β = −0.54, SE = 0.09, p < 0.001 for hypertensive diseases and β = −0.47, SE = 0.09, p < 0.001 for ischemic heart diseases), while higher MET h/week was associated with lower risk of hypertensive and ischemic heart diseases (β = −0.01, SE = 0.00, p < 0.001 and β = −0.01, SE = 0.00, p = 0.005, respectively), thus indicating a potential mediating pathway through reduction in physical activity.

## Discussion

We found that higher genetic liability to LST was associated with more self-reported LST and that the genetic liability to LST also increased the risk of incident CVD, with replicable findings across two independent large populations. The observed effect sizes remained relatively small and attenuated after adjustment, but suggest that the observed adverse health effects may be either directly attributable to genetic effects or mediated by reduced levels of physical activity.

The underlying role of genetics in sedentary behavior and morbidity has so far been understudied.^8^ Evolution has a profound influence on human biology; hence, evolution-based theories provide valuable insights into epidemiology and population health. Evolutionary selection processes may explain why the “desire to be sedentary is in our genes” and also why previously advantageous alleles may currently be disease-causing alleles.^9,10,35^ In our study, the risk of any CVD increased very consistently by 5% per standard deviation increase in the PGS LST. Furthermore, we observed that the risk of CVD differs significantly between those in the highest and lowest deciles of PGS LST. Those with high PGS LST had 21% (19% to 24%) higher risk for any CVD than those with low PGS LST. Those with high PGS LST also indicated higher cumulative disease incidences at different ages, therefore showing lower probability for survival. For example, at the age of 60, those with a high PGS LST had a 7% higher cumulative incidence for any CVD compared to those with a low PGS LST. The associations were statistically independent of SES, BMI, and smoking, although adjustment for these factors attenuated the associations. One explanation for these findings may be genetic pleiotropy.

In genetic pleiotropy, the same genes would affect both LST and CVD. This theory is supported by previously published genetic correlations between LST and several CVD-related phenotypes (0.24 for coronary artery disease, 0.32 for overweight, 0.22 for type 2 diabetes, −0.26 for high-density lipoprotein cholesterol, and 0.27 for triglycerides), indicating shared genetic material.^19^ Previous studies suggest that most robust pleiotropic genes are associated with function of the nervous system, e.g., the DLG4 gene (ENSG00000132535) is associated with synaptic function in the brain, pituitary gland, and retina.^36,37^ Overall, the tissue enrichment analyses suggest that currently known genes associated with LST are expressed mainly in the nervous system, specifically in the brain.^19^ However, more studies are needed to further elucidate the underlying mechanism between genetic predisposition to LST and the onset of CVDs.

Our mediation analysis suggests that there may also be an alternative pathway in which sedentary genes contribute to inadequate levels of physical activity, thereby increasing the risk of incident CVD. Much has been written about the benefits of physical activity on cardiovascular health,^38^ with our findings bringing new perspectives to this discussion. The planning and implementation of public health interventions in different settings (e.g., schools, workplaces, communities) will benefit from an understanding of the inherent biological tendencies of the population.^39^ Considering that genetics contribute to sedentary behavior, effective strategies should emphasize environmental planning that nudges people to be active and/or interventions that significantly nourish the intrinsic reward system.^40^ Moving from sedentary to active behavior may confer health benefits, as the human evolutionary background may also explain why physiological functions operate optimally under regular physical activity.^41^

The observed coefficient of determination and effect size between PGS LST and self-reported LST were small, thus potentially raising concerns how well the score reflects genetic liability to this trait. As there is little prior evidence on how PGSs are associated to sedentary behavior while some data are available for physical activity, we discuss our findings aligned to this construct. Our results are consistent with previous studies in which physical activity PGSs were associated with the corresponding trait and differentiated well the extreme ends of the genetic spectrum, but explain only a small proportion (0.0–1.4%) of the phenotypic variance.^26,42^ This is an interesting contrast and may suggest that current PGSs are not fully able to quantify all the genetic effects they detect. A plausible explanation for the small variances were assumed to be the heterogeneity in phenotype assessment (i.e., physical activity collected through varying methods). However, in our study, we carefully selected the validation data to mimic the phenotype in the base data. Nevertheless, the fraction of variance explained remained small and may indicate other methodological limitations related to polygenic scoring. One of these is the “missing heritability” dilemma. Missing heritability is the difference between the observed SNP heritability and heritability estimates based on twin studies. In our case, this is estimated to be at least 22%^43–45^ and potentially caused by the following: inflated family-based estimates,^46^ relatively small sample sizes of current GWASs,^47^ inadequate accuracy in phenotyping of the trait of interest, and/or the lack of information on less common and rare variants in genotyping arrays used in GWAS studies.

More detailed coverage of the variation in the genome by whole genome sequencing is expected to reduce missing heritability.^48,49^

### Strengths and limitations

This study has several strengths. We used high-performing and genome-wide methods for genetic scoring and showed the scores to be valid with independent data sets. We used the most comprehensive data available and a robust study design, which included a replication cohort. We further tested the robustness of our findings with multiple approaches, with consistent findings supporting our conclusions. We also acknowledge that our results should be interpreted with the following limitations in mind. The base data for PGS LST is based on self-reported LST which may be subject to biases from recall, cultural differences, and social desirability.^50^ Notably, the LST represented screen time activities at the time of the data collection (2011) with limited generalizability to the modern era. The ability to replicate genetic variants related to LST across cohorts remains a challenge, and more quality GWASs regarding sedentary behavior are required.^36^ The data were based on individuals with European ancestry. Hence, replication studies using other populations are required. The FinnGen did not have data related to sedentary behavior or physical activity, and therefore, the mediation analyses could not be conducted in this cohort. The used FinnGen and HUNT may be enriched with CVDs due to participants having had an established contact with healthcare services.

### Conclusions

The findings of this study confirm our hypothesis that higher genetic liability for sedentary behavior is associated with a greater risk of developing CVD, although effect sizes with the current data and methods remain small.

## Data Availability

The utilized FTC subsample data are in the Biobank of the National Institute for Health and Welfare. Data are available for qualified researchers through a standardized application procedure (see the website https://thl.fi/en/web/thl-biobank/for-researchers for details). Access to individual-level genotypes and register data from FinnGen participants can be applied via the Fingenious portal (https://site.fingenious.fi/en/) hosted by the Finnish Biobank Cooperative FinBB (https://finbb.fi/en/). The register data also needs permission from FinData (www.findata.fi). Researchers affiliated with a Norwegian research institution can apply for HUNT data access from the HUNT Research Centre (www.ntnu.edu/hunt) if they have obtained project approval from the Regional Committee for Medical and Health Research Ethics (REC). Researchers not affiliated with a Norwegian research institution should collaborate with and apply through a Norwegian principal investigator. Information on the application and conditions for data access is available online (www.ntnu.edu/hunt/data). The HUNT Databank website provides a detailed overview of the available variables in HUNT (www.ntnu.edu/hunt/databank). Certain data from ancillary HUNT projects may be subject to a time-limited exclusivity provided to the researchers who have financed and conducted the data collection. Biological material is available for analyses, and information on procedures is found on the HUNT Biobank website (www.ntnu.edu/hunt/hunt-biobank). Data from the health registries are not kept by HUNT; instead, linkages between HUNT and registry data must be made for each research project and require that the principal investigator has obtained project-specific approval for such linkage from REC and each registry owner.

## Acknowledgements

We thank the participants and all those who contributed to the data collection from the FTC, FinnGen, and HUNT cohorts without whom this study would not be possible. The detailed list of investigators involved in the FinnGen is available on page 12 of the Supplemental Material.

## Data sharing

The utilized FTC subsample data are in the Biobank of the National Institute for Health and Welfare. Data are available for qualified researchers through a standardized application procedure (see the website https://thl.fi/en/web/thl-biobank/for-researchers for details).

Access to individual-level genotypes and register data from FinnGen participants can be applied via the Fingenious portal (https://site.fingenious.fi/en/) hosted by the Finnish Biobank Cooperative FinBB (https://finbb.fi/en/). The register data also needs permission from FinData (www.findata.fi).

Researchers affiliated with a Norwegian research institution can apply for HUNT data access from the HUNT Research Centre (www.ntnu.edu/hunt) if they have obtained project approval from the Regional Committee for Medical and Health Research Ethics (REC). Researchers not affiliated with a Norwegian research institution should collaborate with and apply through a Norwegian principal investigator. Information on the application and conditions for data access is available online (www.ntnu.edu/hunt/data). The HUNT Databank website provides a detailed overview of the available variables in HUNT (www.ntnu.edu/hunt/databank). Certain data from ancillary HUNT projects may be subject to a time-limited exclusivity provided to the researchers who have financed and conducted the data collection. Biological material is available for analyses, and information on procedures is found on the HUNT Biobank website (www.ntnu.edu/hunt/hunt-biobank). Data from the health registries are not kept by HUNT; instead, linkages between HUNT and registry data must be made for each research project and require that the principal investigator has obtained project-specific approval for such linkage from REC and each registry owner.

## Contributorship

LJ and ES conceptualized the research question, study design, and statistical analysis. KK, UE, and JK made significant contributions to the final analysis design. LJ prepossessed the polygenic score data under the supervision of TP and implemented the scores to the FTC, FinnGen, and HUNT study cohorts with the help of TP, KK, and NT. LJ performed the statistical modelling under supervision of ES and with consultation from NT. JK and TP participated in FTC data collection and management, while AB, MK, KØ, and UW to HUNT data. LJ drafted the first version of the manuscript, and all authors contributed significantly to the writing, critical interpretation of the findings, and revising the manuscript. ES acquired funding for the study. The corresponding author attests that all listed authors meet authorship criteria and that no others meeting the criteria have been omitted.

