## Supplemental Material for "Genetic liability to sedentary behavior increases the risk of cardiovascular disease incidence: Evidence from the FinnGen cohort with 293,250 individuals"

### Supplemental Methods

#### Ethics in the FinnGen

Recruitment protocols followed the biobank protocols approved by Fimea. The Coordinating Ethics Committee of the Hospital District of Helsinki and Uusimaa (HUS) approved the FinnGen study protocol (number HUS/990/2017). The FinnGen study protocol (number HUS/990/2017) was approved by the Coordinating Ethics Committee of the Hospital District of Helsinki and Uusimaa (HUS). The FinnGen study is approved by the THL (approval number THL/2031/6.02.00/2017, amendments THL/1101/5.05.00/2017, THL/341/6.02.00/2018, THL/2222/6.02.00/2018, THL/283/6.02.00/2019 and THL/1721/5.05.00/2019), the Digital and Population Data Service Agency (VRK43431/2017-3, VRK/6909/2018-3 and VRK/4415/2019-3), the Social Insurance Institution (KELA) (KELA 58/522/2017, KELA 131/522/2018, KELA 70/522/2019 and KELA 98/522/2019) and Statistics Finland (TK-53-1041-17).

The Biobank Access Decisions for FinnGen samples and data utilized in FinnGen include the following datasets: THL Biobank BB2017_55, BB2017_111, BB2018_19, BB_2018_34, BB_2018_67, BB2018_71, BB2019_7, BB2019_8 and BB2019_26; Finnish Red Cross Blood Service Biobank 7.12.2017; Helsinki Biobank HUS/359/2017; Auria Biobank AB17-5154; Biobank Borealis of Northern Finland_2017_1013; Biobank of Eastern Finland 1186/2018; Finnish Clinical Biobank Tampere MH0004; Central Finland Biobank 1-2017; and Terveystalo Biobank STB 2018001.

#### Genotyping in the Finnish Twin Cohort (FTC).

In the FTC, Chip genotyping was done using Illumina Human610-Quad v1.0 B, Human670-QuadCustom v1.0 A, Illumina HumanCoreExome- (12 v1.0 B, 12 v1.1 A, 24 v1.0 A, 24 v1.1 A, 24 v1.2A) and Affymetrix FinnGen Axiom arrays. Genotype quality control was done in three batches (batch1: 610k+670k chip, batch2: HumanCoreExome and batch3: Affymetrix chip genotypes) with removing variants with call rate below 97,5% (batch1 and batch3) and 95% (batch2), removing samples with call rate below 98% (batch1) or 95% (batch2 and batch3), removing variants with its minor allele frequency below 1% and Hardy-Weinberg Equilibrium p-value lower than 1e-06. Also samples from all batches with heterozygosity test method-of-moments F coefficient estimate value below -0.03 or higher than 0.05 (batch1 and batch2) or ±4SD from the mean (batch3) were removed along with the samples which failed sex check or were among the MDS principal component analysis outliers. Total amount of genotyped autosomal variants after QC were 475526 (batch1), 239894 (batch2) and 388673 (batch3). We then performed pre-phasing using Eagle v2.3,[1] and imputation with Minimac3 v2.0.1 using University of Michigan Imputation Server.[2] Genotypes of all batches were imputed to Haplotype Reference Consortium release 1.1 reference panel.[3]

### Supplemental Tables

Table S1. Descriptives of the cohorts included in the polygenic score base data previously published by Wang et al (2022).

| COHORTS | | Age (years) | BMI (kg/m2) | Leisure screen time (h/day) |
|---|---|---|---|---|
| **ALSPAC-MOTHERS** | women | 36.4 (4.5) | 24.3 (4.2) | 2.6 (1.6) |
| **ALSPAC-OFFSPRING** | men | 16.7 (0.3) | 20.9 (3.3) | 1.8 (1.0) |
| | women | 16.7 (0.2) | 21.6 (3.5) | 1.7 (1.0) |
| **B58C-WTCCC2** | men | 45.1 (0.4) | 27.7 (4.2) | 2.2 (1.1) |
| | women | 45.1 (0.4) | 26.8 (5.4) | 2.2 (1.2) |
| **B58C- T1DGC** | men | 45.3 (0.3) | 28.0 (4.1) | 2.2 (1.1) |
| | women | 45.3 (0.3) | 27.0 (5.6) | 2.2 (1.1) |
| **CARDIA** | men | 30.6 (3.3) | 25.6 (3.8) | 1.8 (1.6) |
| | women | 30.5 (3.4) | 24.2 (4.8) | 1.6 (1.4) |
| **COLAUS ROUND 2** | men | 57.9 (10.6) | 26.8 (4.0) | 1.1 (1.1) |
| | women | 58.7 (10.4) | 25.4 (4.9) | 0.9 (1.1) |
| | omni men | 49.6 (19.5) | 26.7 (4.7) | 1.8 (1.3) |
| | omni women | 50.2 (20.4) | 26.5 (5.5) | 1.8 (1.3) |
| **EPIC-NORFOLK** | men | 59.6 (9.2) | 26.5 (3.2) | 3.1 (1.4) |
| | women | 58.8 (9.3) | 26.1 (4.2) | 3.1 (1.4) |
| **FAMHS** | men | 52.0 (13.9) | 28.0 (4.6) | 2.2 (1.5) |
| | women | 52.4 (13.4) | 27.5 (6.2) | 2.1 (1.5) |
| **FENLAND-OMICS** | men | 48.8 (7.4) | 27.4 (4.2) | 2.7 (1.4) |
| | women | 48.9 (7.3) | 26.6 (5.4) | 2.7 (1.5) |
| **GENOA** | men | 59.5 (10.1) | 30.8 (5.2) | 2.4 (1.5) |
| | women | 58.6 (10.2) | 30.8 (7.0) | 2.4 (1.5) |
| **GOOD** | men | 18.9 (0.6) | 22.4 (3.2) | 3.2 (2.2) |
| **GRAPHIC** | men | 53.8 (4.2) | 27.8 (3.9) | 5.7 (1.5) |
| | women | 51.9 (4.4) | 27.1 (4.6) | 5.7 (1.6) |
| **HPFSAFFY** | case men | 55.9 (8.6) | 27.1 (3.9) | 1.6 (1.3) |

| | | | | |
|---|---|---|---|---|
| | control men | 55.9 (8.5) | 25.0 (2.8) | 1.5 (1.2) |
| **HPFSILLUMINA** | case men | 53.5 (8.9) | 25.2 (2.9) | 1.4 (1.2) |
| | control men | 54.8 (8.5) | 25.5 (2.9) | 1.5 (1.2) |
| **HPFSOMNI** | case men | 54.7 (8.8) | 26.2 (3.0) | 1.5 (1.2) |
| | control men | 54.5 (8.8) | 25.3 (2.9) | 1.4 (1.2) |
| **HRS** | men | 67.7 (10.1) | 29.2 (4.9) | 3.1 (1.8) |
| | women | 67.0 (11.2) | 28.8 (6.4) | 3.0 (1.9) |
| **MESA** | men | 62.8 (10.1) | 28.0 (4.2) | 1.9 (1.4) |
| | women | 62.7 (10.2) | 27.7 (5.9) | 2.0 (1.5) |
| **NESDA** | men | 44.3 (12.3) | 26.5 (4.7) | 3.0 (1.9) |
| | women | 41.5 (12.9) | 25.4 (5.3) | 3.0 (1.8) |
| **NHSAFFY** | case women | 54.5 (6.8) | 28.7 (5.7) | 2.1 (1.9) |
| | control women | 54.5 (6.8) | 24.9 (4.5) | 1.9 (1.6) |
| **NHSILLUMINA** | case women | 54.5 (6.7) | 25.0 (4.5) | 1.9 (1.7) |
| | control women | 54.6 (6.5) | 24.6 (4.4) | 1.9 (1.6) |
| **NHSOMNI** | case women | 54.4 (6.7) | 26.6 (5.8) | 1.9 (1.8) |
| | control women | 54.7 (6.6) | 25.1 (4.5) | 1.9 (1.6) |
| **UK BIOBANK** | men | 57.0 (8.1) | 27.8 (4.2) | 3.6 (1.9) |
| | women | 56.6 (8.0) | 27.0 (5.1) | 4.0 (2.0) |
| **WGHS** | women | 54.2 (7.1) | 25.9 (5.0) | 1.8 (2.0) |
| **YFS** | men | 32.7 (5.7) | 26.0 (4.2) | 2.0 (1.2) |
| | women | 32.7 (5.7) | 24.7 (4.8) | 1.8 (1.1) |
| **ALL** | **MEAN** | **49.5 (7.6)** | **26.4 (4.5)** | **2.3 (1.5)** |

Data are means and (standard deviations) originally described by Wang et al (2022) in their Supplementary Table 1. ALSPAC, Avon Longitudinal Study of Parents and Children; B58C, the 1958 Birth Cohort; CARDIA, the Coronary Artery Risk Development in Young Adults Study; CoLAUS, a population-based cohort study in Lausanne; EPIC-Norfolk, European Prospective Investigation into Cancer, Norfolk study; FAMHS, Family Heart Study; Fenland-OMICS, The Fenland Study; GENOA, Genetic Epidemiology Network of Arteriosclerosis; GOOD, The Gothenburg Osteoporosis and Obesity Determinants study; GRAPHIC, The GRAPHIC Study; HPFSaffy, Health Professional Follow-up Study; HPFSillumina, Health Professional Follow-up Study; HPFSomni, Health Professional Follow-up Study; HRS, Health and retirement study; MESA, Multi-Ethnic Study of Atherosclerosis; NESDA, The Netherlands Study of Depression and Anxiety; NHSaffy, Nurse Health Study; NHSillumina, Nurse Health Study; NHSomni, Nurse Health Study; UK BioBank; WGHS, Women's Genome Health Study; YFS, The Young Finns Study. All data from Wang et al (2022).[4]

Table S2. Descriptives of the Finnish Twin Cohort participants with polygenic scores for leisure screen time

| **Characteristic** | **N** | **Mean (SD) or %** |
|---|---|---|
| Age (years) | 2,689 | 60.5 (3.7) |
| Female (%) | 1,540 | 54.7 |
| Educational attainment | | |
| High | 546 | 21.6 |
| Middle | 1,293 | 51.3 |
| Low | 683 | 27.1 |
| BMI ($kg/m^2$) | 2,638 | 26.5 (4.4) |
| LST (h/day) | 2,689 | 3.9 (1.1) |

Educational attainment was based on the Finnish education system: High, at least upper secondary degree; Middle, basic education and additional studies; Low, basic education degree at most; BMI, body mass index; LST, self-reported leisure screen time.

Table S3. Association between polygenic score for leisure screen time and self-reported leisure screen time in the Finnish Twin Cohort.

| | | **LST (h/day)** | |
|---|---|---|---|
| Models | N | β | 95 % CI |
| Crude model | 2,689 | **0.09** | **0.05 to 0.14** |
| Model 1 | 2,689 | **0.09** | **0.05 to 0.13** |
| Model 2 | 2,478 | **0.08** | **0.04 to 0.13** |

LST, Leisure screen time in hours per day; Results indicate increments in self-reported leisure screen time (hours per day) per one standard deviation increase in polygenic score for leisure screen time (PGS LST). Crude model: PGS LST standardized score, ten principal components of ancestry; Model 1: Additionally, age and sex; Model 2: Additionally, educational attainment and body mass index. Statistically significant associations highlighted with **bold.**

Table S4. Individuals with high and low polygenic score for leisure screen time and their self-reported leisure screen time in the Finnish Twin Cohort

| | **PGS LST percentile** | | |
|---|---|---|---|
| | < 10th | > 90th | P for difference |
| LST (h/day) | 3.7 (1.1) | 4.1 (1.0) | **<0.001** |

LST, Leisure screen time in hours per day; PGS LST percentile, genetic liability for leisure screen time estimated with a polygenic score; < $10^{th}$, individuals in the lowest decile in the genetic spectrum; > $90^{th}$, individuals in the highest decile in the genetic spectrum. Difference between the groups is evaluated with linear regression adjusted for ten principal components of ancestry, age, sex, educational attainment and body mass index. Statistically significant differences highlighted with **bold.**

Table S5. FinnGen participants with high and low polygenic score for leisure screen time and incident CVD

| Cardiovascular diseases | Cases/N | HR (95% CI) |
|---|---|---|
| **Any CVD** | | |
| <10th PGS LST | 16,234/29,325 | 1.00 (Reference) |
| >90th PGS LST | 17,483/29,325 | **1.21 (1.19 to 1.24)** |
| **Hypertensive diseases** | | |
| <10th PGS LST | 9,804/32,289 | 1.00 (Reference) |
| >90th PGS LST | 11,753/32,289 | **1.35 (1.31 to 1.39)** |
| **Ischemic heart diseases** | | |
| <10th PGS LST | 6,067/33,302 | 1.00 (Reference) |
| >90th PGS LST | 6,864/33,301 | **1.26 (1.22 to 1.31)** |
| **Cerebrovascular diseases** | | |
| <10th PGS LST | 3,294/33,273 | 1.00 (Reference) |
| >90th PGS LST | 3,584/33,273 | **1.19 (1.13 to 1.24)** |

Values are hazard ratios (HR) and 95% confidence intervals (CI) in high polygenic score group for leisure-screen time (PGS LST) compared to low PGS LST group. In high PGS LST the sample-specific PGS LST percentile was >90$^{th}$ while in low PGS LST <10$^{th}$. We used age as the time scale, and ten principal components of ancestry, sex, and genotyping batch as covariates; CVD, cardiovascular disease. Statistically significant associations highlighted with **bold**.

Table S6. HUNT participants with high and low polygenic score for leisure screen time and incident CVD

| Cardiovascular diseases | Cases/N | HR (95% CI) |
|---|---|---|
| **Any CVD** | | |
| <10th PGS LST | 3,070/3,529 | 1.00 (Reference) |
| >90th PGS LST | 2985/3,528 | **1.16 (1.10 to 1.22)** |
| **Hypertensive diseases** | | |
| <10th PGS LST | 1,430/3,634 | 1.00 (Reference) |
| >90th PGS LST | 1,545/3,633 | **1.28 (1.19 to 1.37)** |
| **Ischemic heart diseases** | | |
| <10th PGS LST | 1,067/3,638 | 1.00 (Reference) |
| >90th PGS LST | 1,131/3,737 | **1.26 (1.16 to 1.37)** |
| **Cerebrovascular diseases** | | |
| <10th PGS LST | 659/3639 | 1.00 (Reference) |
| >90th PGS LST | 603/3638 | 1.08 (0.96 to 1.20) |

Values are hazard ratios (HR) and 95% confidence intervals (CI) in high polygenic score group for leisure-screen time (PGS LST) compared to low PGS LST group. In high PGS LST the sample-specific PGS LST percentile was >90$^{th}$ while in low PGS LST <10$^{th}$. We used age as the time scale, and ten principal components of ancestry, sex, and genotyping batch as covariates; CVD, cardiovascular disease. Statistically significant associations highlighted with **bold**.

Table S7. Cumulative incidence estimates at selected ages in the FinnGen cohort

| **Cumulative incidence** | **FinnGen** | | | | **P-value** |
|---|---|---|---|---|---|
| | **<10th** | **95% CI** | **>90th** | **95% CI** | |
| **Any CVD** | | | | | |
| 60 y | 0.37 | 0.36 to 0.38 | 0.44 | 0.43 to 0.45 | |
| 80 y | 0.82 | 0.81 to 0.82 | 0.87 | 0.86 to 0.88 | **<0.001** |
| **Hypertensive diseases** | | | | | |
| 60 y | 0.17 | 0.17 to 0.18 | 0.24 | 0.24 to 0.25 | |
| 80 y | 0.49 | 0.48 to 0.50 | 0.59 | 0.59 to 0.60 | **<0.001** |
| **Ischemic heart disease** | | | | | |
| 60 y | 0.06 | 0.06 to 0.07 | 0.09 | 0.08 to 0.09 | |
| 80 y | 0.31 | 0.30 to 0.32 | 0.38 | 0.37 to 0.39 | **<0.001** |
| **Cerebrovascular diseases** | | | | | |
| 60 y | 0.03 | 0.03 to 0.03 | 0.04 | 0.04 to 0.04 | |
| 80 y | 0.17 | 0.17 to 0.18 | 0.20 | 0.20 to 0.21 | **<0.001** |

Cumulative incidence estimates for low (<10th percentile) and high (>90th percentile) polygenic score for leisure screen time (PGS LST) groups in the FinnGen by the function of age. Values are from Cox proportional hazards models and adjusted for ten principal components, sex and genotyping batch. CVD, cardiovascular disease. Statistically significant differences between groups highlighted with **bold** analysed with Gray's test.

Table S8. Cumulative incidence estimates for competing risks at selected ages in the FinnGen cohort

| **Cumulative incidence** | **FinnGen** | | | | **P-value** |
|---|---|---|---|---|---|
| | **<10th** | **95% CI** | **>90th** | **95% CI** | |
| **Hypertensive diseases** with ischemic heart diseases and cerebrovascular diseases as competing risks | | | | | |
| 60 y | 0.05 | 0.05 to 0.06 | 0.09 | 0.08 to 0.09 | |
| 80 y | 0.40 | 0.39 to 0.41 | 0.48 | 0.48 to 0.49 | **<0.001** |
| **Ischemic heart disease** with cerebrovascular diseases and hypertensive diseases as competing risks | | | | | |
| 60 y | 0.01 | 0.01 to 0.02 | 0.02 | 0.02 to 0.02 | |
| 80 y | 0.16 | 0.15 to 0.16 | 0.17 | 0.16 to 0.18 | **0.003** |
| **Cerebrovascular diseases** with hypertensive diseases and ischemic heart diseases as competing risks | | | | | |
| 60 y | 0.01 | 0.01 to 0.01 | 0.01 | 0.01 to 0.01 | |
| 80 y | 0.07 | 0.07 to 0.08 | 0.08 | 0.07 to 0.08 | 0.700 |

Cumulative incidence estimates for low (<10th percentile) and high (>90th percentile) polygenic score for leisure screen time (PGS LST) groups in the FinnGen by the function of age. Values are from competing risk cumulative incidence estimates. CVD, cardiovascular disease. P-value, difference between low and high PGS LST groups; Statistically significant differences between groups highlighted with **bold** analysed with Gray's test.

Table S9. Sensitivity analyses for associations of polygenic score for leisure screen with CVD incidence with baseline from DNA sampling

| Cardiovascular diseases | Cases/N | Incidence/ 10,000 person-years | HR (95% CI) |
|---|---|---|---|
| **Any CVD** | | | |
| FinnGen | 41,295/142,026 | 193.9 | **1.05 (1.04 to 1.06)** |
| HUNT | 8,063/10,070 | 609.6 | 1.01 (0.98 to 1.03) |
| **Hypertensive diseases** | | | |
| FinnGen | 31,930/216,121 | 97.7 | **1.07 (1.06 to 1.08)** |
| HUNT | 6,240/15,642 | 259.3 | **1.03 (1.00 to 1.05)** |
| **Ischemic heart disease** | | | |
| FinnGen | 25,879/260,552 | 59.1 | **1.04 (1.02 to 1.05)** |
| HUNT | 2,761/15,778 | 107.6 | **1.04 (1.00 to 1.08)** |
| **Cerebrovascular diseases** | | | |
| FinnGen | 17,014/281,668 | 32.4 | **1.04 (1.03 to 1.06)** |
| HUNT | 2,058/ 17,904 | 61.0 | 1.01 (0.96 to 1.05) |

In sensitivity analyses the baseline was set to the individual DNA sampling date. Values are Hazard Ratios (HR) and 95% Confidence intervals (95% CI) per one standard deviation increase in the polygenic score for leisure screen time; Values are adjusted with ten first principle components of ancestry, sex and batch; CVD, cardiovascular disease; The mean follow-up was 6.0 years (from minimum of 0.0 to maximum of 47.5 years) and a total of 857,702 person-years with any CVDs in the FinnGen and the mean of 5.0 years (min 0.1, max 11.1 years) and a total of 51,712 person-years in the HUNT. Incidence values are age-standardised according to the European Standard Population to enable comparison between FinnGen and HUNT. Statistically significant associations highlighted with **bold.**

### Supplemental Figures

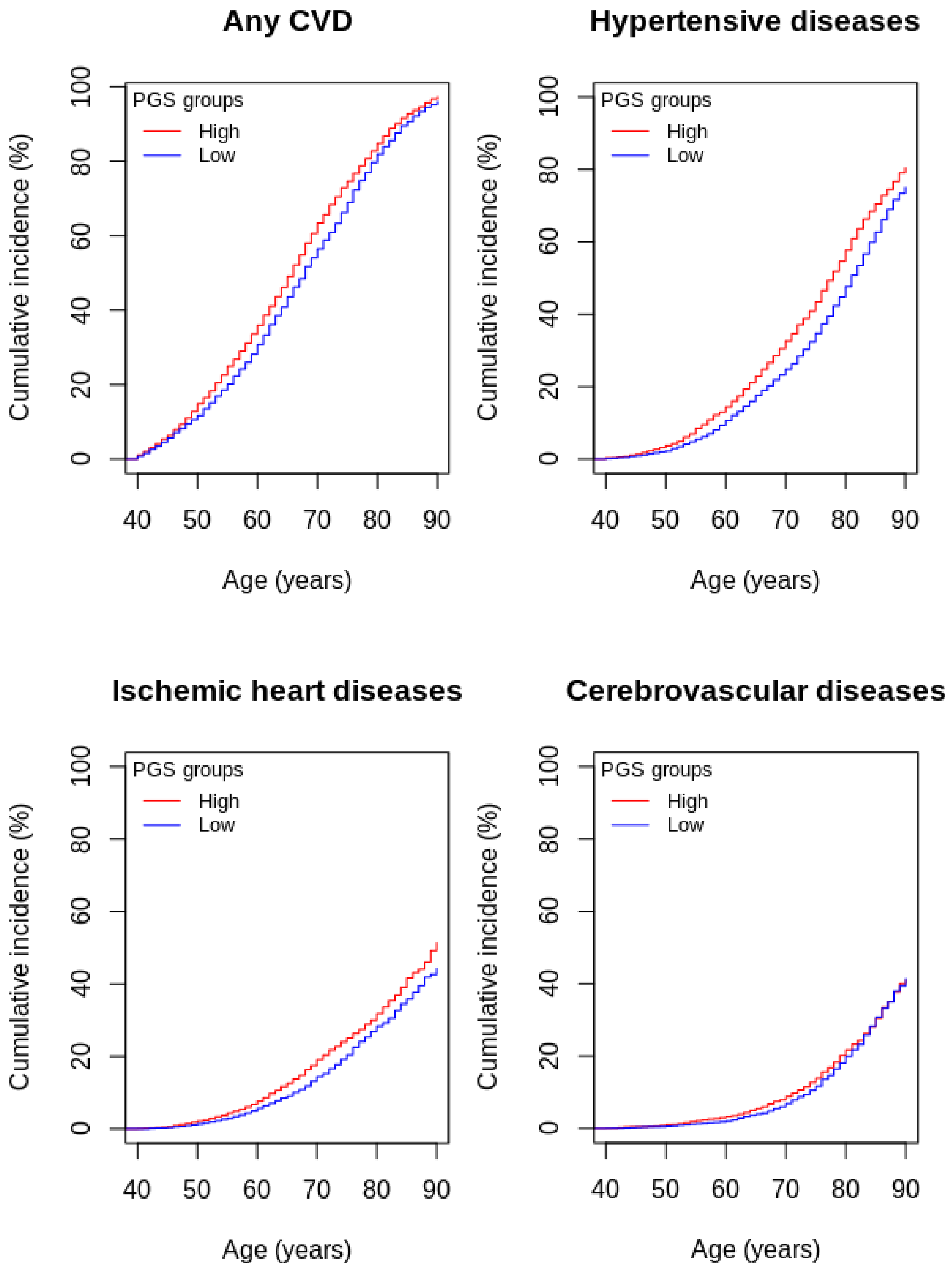


Figure S1. Cumulative incidence curves for high (>90th percentile [red]) and low (<10th percentile [blue]) PGS LST groups in HUNT based on Cox proportional hazards models and adjusted for ten principal components, sex, and genotyping batch.

| Cardiovascular disease | Cases/N | HR (95% CI) |
|---|---|---|
| **Any CVD** | | |
| Model 1 | 17,362/20,383 | 1.03 (1.01 to 1.04) |
| Model 2 + SES | 17,362/20,383 | 1.03 (1.02 to 1.05) |
| Model 3 + BMI | 17,362/20,383 | 1.02 (1.01 to 1.04) |
| Model 4 + Smoking | 17,362/20,383 | 1.02 (1.00 to 1.03) |
| **Hypertensive diseases** | | |
| Model 1 | 8,987/21,121 | 1.06 (1.04 to 1.09) |
| Model 2 + SES | 8,987/21,121 | 1.07 (1.04 to 1.09) |
| Model 3 + BMI | 8,987/21,121 | 1.04 (1.02 to 1.06) |
| Model 4 + Smoking | 8,987/21,121 | 1.03 (1.01 to 1.05) |
| **Ischemic heart diseases** | | |
| Model 1 | 5,647/21,148 | 1.05 (1.02 to 1.08) |
| Model 2 + SES | 5,647/21,148 | 1.05 (1.02 to 1.08) |
| Model 3 + BMI | 5,647/21,148 | 1.04 (1.01 to 1.07) |
| Model 4 + Smoking | 5,647/21,148 | 1.03 (1.00 to 1.06) |
| **Cerebrovascular diseases** | | |
| Model 1 | 2,786/21,156 | 1.05 (1.01 to 1.09) |
| Model 2 + SES | 2,786/21,156 | 1.05 (1.01 to 1.09) |
| Model 3 + BMI | 2,786/21,156 | 1.05 (1.01 to 1.09) |
| Model 4 + Smoking | 2,786/21,156 | 1.03 (0.99 to 1.07) |

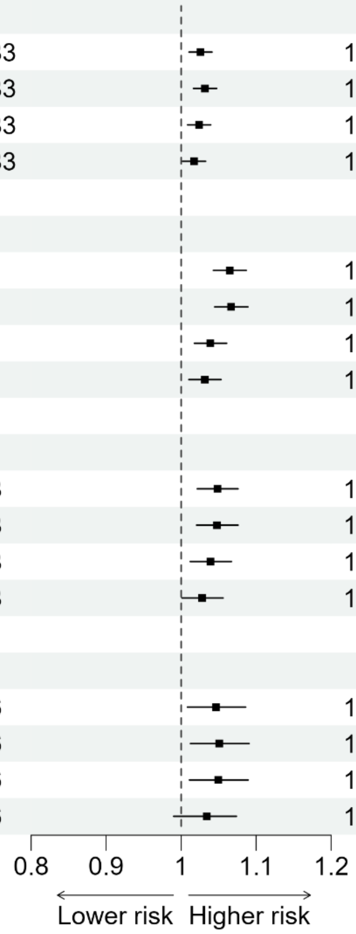


Figure S2. Associations between polygenic score for leisure screen time and incident cardiovascular disease in HUNT adjusted with covariates. Values are hazard ratios (HR) and 95% confidence intervals (CI) per one standard deviation increase in the polygenic score for leisure screen time. We used age as the time scale. Model 1: Polygenic score for leisure screen time, ten principal components of ancestry, sex, and genotyping batch; Model 2, additionally socioeconomic status (SES); Model 3, additionally body mass index (BMI); Model 4: additionally smoking status. CVD, cardiovascular disease.

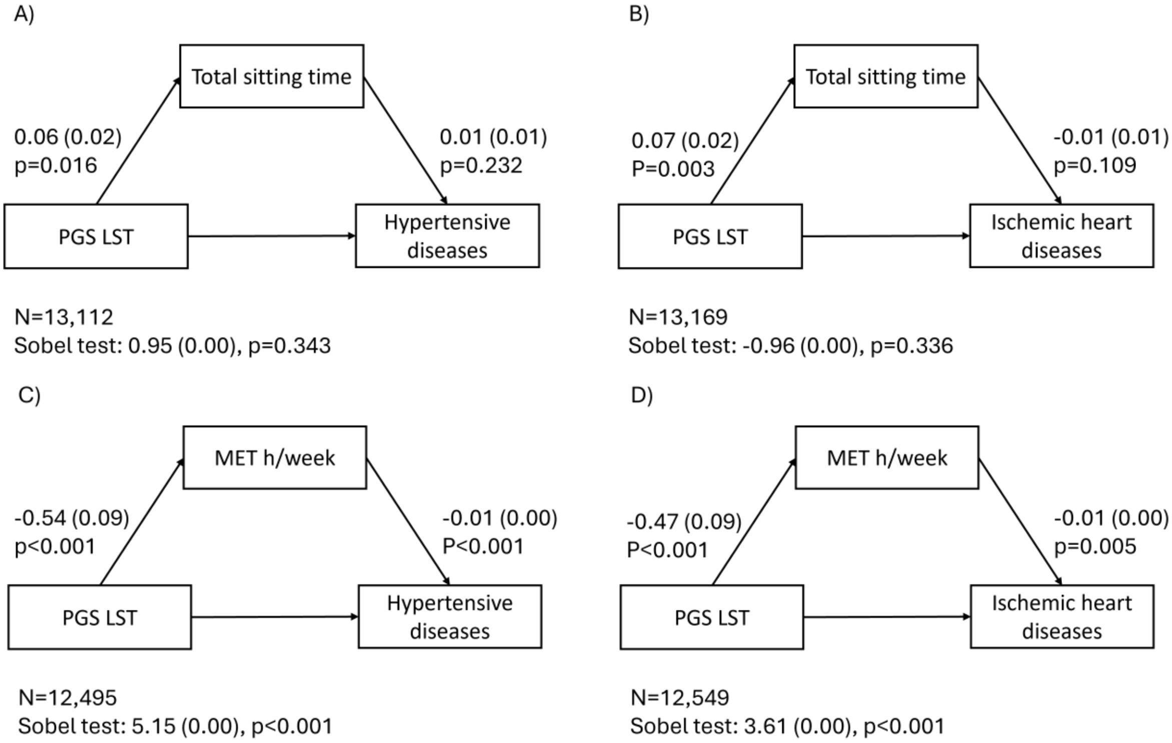


Figure S3. Exploratory mediation analysis using the product of coefficients approach to evaluate potential mediation pathways. Panels A and B show associations for total sitting time (h/day) and C and D for metabolic equivalent (MET) hours per week (h/week). Values are unstandardized regression coefficients (standard errors) and p-values.

### FinnGen

| Full Name | Affiliation | E-mail | Role 1 | Role 2 |
|---|---|---|---|---|
| Aarno Palotie | Institute for Molecular Medicine Finland (FIMM), HiLIFE, University of Helsinki, Helsinki, | | Steering Committee | Steering Committee |
| Mark Daly | Institute for Molecular Medicine Finland (FIMM), HiLIFE, University of Helsinki, Helsinki, | | Steering Committee | Steering Committee |
| Bridget Riley-Gills | Abbvie, Chicago, IL, United States | | Steering Committee | Pharmaceutical companies |
| Howard Jacob | Abbvie, Chicago, IL, United States | | Steering Committee | Pharmaceutical companies |
| Dirk Paul | Astra Zeneca, Cambridge, United Kingdom | | Steering Committee | Pharmaceutical companies |
| Slavé Petrovski | Astra Zeneca, Cambridge, United Kingdom | | Steering Committee | Pharmaceutical companies |
| Heiko Runz | Biogen, Cambridge, MA, United States | | Steering Committee | Pharmaceutical companies |
| Sally John | Biogen, Cambridge, MA, United States | | Steering Committee | Pharmaceutical companies |
| George Okafo | Boehringer Ingelheim, Ingelheim am Rhein, Germany | | Steering Committee | Pharmaceutical companies |
| Robert Plenge | Bristol Myers Squibb, New York, NY, United States | | Steering Committee | Pharmaceutical companies |
| Joseph Maranville | Bristol Myers Squibb, New York, NY, United States | | Steering Committee | Pharmaceutical companies |
| Mark McCarthy | Genentech, San Francisco, CA, United States | | Steering Committee | Pharmaceutical companies |
| Margaret G. Ehm | GlaxoSmithKline, Collegeville, PA, United States | | Steering Committee | Pharmaceutical companies |
| Kirsi Auro | GlaxoSmithKline, Espoo, Finland | | Steering Committee | Pharmaceutical companies |
| Simonne Longerich | Merck, Kenilworth, NJ, United States | | Steering Committee | Pharmaceutical companies |
| Anders Mälarstig | Pfizer, New York, NY, United States | | Steering Committee | Pharmaceutical companies |
| Katherine Klinger | Translational Sciences, Sanofi R&D, Framingham, MA, USA | | Steering Committee | Pharmaceutical companies |
| Clement Chatelain | Translational Sciences, Sanofi R&D, Framingham, MA, USA | | Steering Committee | Pharmaceutical companies |
| Matthias Gossel | Translational Sciences, Sanofi R&D, Framingham, MA, USA | | Steering Committee | Pharmaceutical companies |
| Karol Estrada | Maze Therapeutics, San Francisco, CA, United States | | Steering Committee | Pharmaceutical companies |
| Robert Graham | Maze Therapeutics, San Francisco, CA, United States | | Steering Committee | Pharmaceutical companies |
| Dawn Waterworth | Janssen Research & Development, LLC, Spring House, PA, United States | | Steering Committee | Pharmaceutical companies |
| Chris O´Donnell | Novartis Institutes for BioMedical Research, Cambridge, MA, United States | | Steering Committee | Pharmaceutical companies |
| Nicole Renaud | Novartis Institutes for BioMedical Research, Cambridge, MA, United States | | Steering Committee | Pharmaceutical companies |
| Tomi P. Mäkelä | HiLIFE, University of Helsinki, Finland, Finland | | Steering Committee | University of Helsinki & Biobanks |
| Jaakko Kaprio | Institute for Molecular Medicine Finland (FIMM), HiLIFE, University of Helsinki, Helsinki, | | Steering Committee | University of Helsinki & Biobanks |
| Petri Virolainen | Auria Biobank / University of Turku / Hospital District of Southwest Finland, Turku, | | Steering Committee | University of Helsinki & Biobanks |
| Antti Hakanen | Auria Biobank / University of Turku / Hospital District of Southwest Finland, Turku, | | Steering Committee | University of Helsinki & Biobanks |
| Terhi Kilpi | THL Biobank / Finnish Institute for Health and Welfare (THL), Helsinki, Finland | | Steering Committee | University of Helsinki & Biobanks |
| Markus Perola | THL Biobank / Finnish Institute for Health and Welfare (THL), Helsinki, Finland | | Steering Committee | University of Helsinki & Biobanks |
| Jukka Partanen | Finnish Red Cross Blood Service / Finnish Hematology Registry and Clinical Biobank, | | Steering Committee | University of Helsinki & Biobanks |
| Anne Pitkäranta | Helsinki Biobank / Helsinki University and Hospital District of Helsinki and Uusimaa, | | Steering Committee | University of Helsinki & Biobanks |
| Taneli Raivio | Helsinki Biobank / Helsinki University and Hospital District of Helsinki and Uusimaa, | | Steering Committee | University of Helsinki & Biobanks |
| Jani Tikkanen | Northern Finland Biobank Borealis / University of Oulu / Northern Ostrobothnia Hospital | | Steering Committee | University of Helsinki & Biobanks |
| Raisa Serpi | Northern Finland Biobank Borealis / University of Oulu / Northern Ostrobothnia Hospital | | Steering Committee | University of Helsinki & Biobanks |
| Tarja Laitinen | Finnish Clinical Biobank Tampere / University of Tampere / Pirkanmaa Hospital | | Steering Committee | University of Helsinki & Biobanks |
| Veli-Matti Kosma | Biobank of Eastern Finland / University of Eastern Finland / Northern Savo Hospital | | Steering Committee | University of Helsinki & Biobanks |
| Jari Laukkanen | Central Finland Biobank / University of Jyväskylä / Central Finland Health Care District, | | Steering Committee | University of Helsinki & Biobanks |
| Marco Hautalahti | FINBB - Finnish biobank cooperative | | Steering Committee | University of Helsinki & Biobanks |
| Outi Tuovila | Business Finland, Helsinki, Finland | | Steering Committee | Other Experts/ Non-Voting Members |
| Raimo Pakkanen | Business Finland, Helsinki, Finland | | Steering Committee | Other Experts/ Non-Voting Members |
| Jeffrey Waring | Abbvie, Chicago, IL, United States | | Scientific Committee | Pharmaceutical companies |
| Bridget Riley-Gillis | Abbvie, Chicago, IL, United States | | Scientific Committee | Pharmaceutical companies |
| Fedik Rahimov | Abbvie, Chicago, IL, United States | | Scientific Committee | Pharmaceutical companies |
| Ioanna Tachmazidou | Astra Zeneca, Cambridge, United Kingdom | | Scientific Committee | Pharmaceutical companies |
| Chia-Yen Chen | Biogen, Cambridge, MA, United States | | Scientific Committee | Pharmaceutical companies |
| Heiko Runz | Biogen, Cambridge, MA, United States | | Scientific Committee | Pharmaceutical companies |
| Zhihao Ding | Boehringer Ingelheim, Ingelheim am Rhein, Germany | | Scientific Committee | Pharmaceutical companies |
| Marc Jung | Boehringer Ingelheim, Ingelheim am Rhein, Germany | | Scientific Committee | Pharmaceutical companies |
| Hanati Tuoken | Boehringer Ingelheim, Ingelheim am Rhein, Germany | | Scientific Committee | Pharmaceutical companies |
| Shameek Biswas | Bristol Myers Squibb, New York, NY, United States | | Scientific Committee | Pharmaceutical companies |
| Rion Pendergrass | Genentech, San Francisco, CA, United States | | Scientific Committee | Pharmaceutical companies |
| Margaret G. Ehm | GlaxoSmithKline, Collegeville, PA, United States | | Scientific Committee | Pharmaceutical companies |
| David Pulford | GlaxoSmithKline, Stevenage, United Kingdom | | Scientific Committee | Pharmaceutical companies |
| Neha Raghavan | Merck, Kenilworth, NJ, United States | | Scientific Committee | Pharmaceutical companies |
| Adriana Huertas-Vazque | Merck, Kenilworth, NJ, United States | | Scientific Committee | Pharmaceutical companies |
| Jae-Hoon Sul | Merck, Kenilworth, NJ, United States | | Scientific Committee | Pharmaceutical companies |
| Anders Mälarstig | Pfizer, New York, NY, United States | | Scientific Committee | Pharmaceutical companies |
| Xinli Hu | Pfizer, New York, NY, United States | | Scientific Committee | Pharmaceutical companies |
| Åsa Hedman | Pfizer, New York, NY, United States | | Scientific Committee | Pharmaceutical companies |
| Katherine Klinger | Translational Sciences, Sanofi R&D, Framingham, MA, USA | | Scientific Committee | Pharmaceutical companies |
| Robert Graham | Maze Therapeutics, San Francisco, CA, United States | | Scientific Committee | Pharmaceutical companies |
| Manuel Rivas | Maze Therapeutics, San Francisco, CA, United States | | Scientific Committee | Pharmaceutical companies |
| Dawn Waterworth | Janssen Research & Development, LLC, Spring House, PA, United States | | Scientific Committee | Pharmaceutical companies |
| Nicole Renaud | Novartis Institutes for BioMedical Research, Cambridge, MA, United States | | Scientific Committee | Pharmaceutical companies |
| Ma´en Obeidat | Novartis Institutes for BioMedical Research, Cambridge, MA, United States | | Scientific Committee | Pharmaceutical companies |
| Jonathan Chung | Novartis Institutes for BioMedical Research, Cambridge, MA, United States | | Scientific Committee | Pharmaceutical companies |
| Jonas Zierer | Novartis Institutes for BioMedical Research, Cambridge, MA, United States | | Scientific Committee | Pharmaceutical companies |
| Mari Niemi | Novartis Institutes for BioMedical Research, Cambridge, MA, United States | | Scientific Committee | Pharmaceutical companies |
| Samuli Ripatti | Institute for Molecular Medicine Finland (FIMM), HiLIFE, University of Helsinki, Helsinl | | Scientific Committee | University of Helsinki & Biobanks |
| Johanna Schleutker | Auria Biobank / Univ. of Turku / Hospital District of Southwest Finland, Turku, Finland | | Scientific Committee | University of Helsinki & Biobanks |
| Markus Perola | THL Biobank / Finnish Institute for Health and Welfare (THL), Helsinki, Finland | | Scientific Committee | University of Helsinki & Biobanks |
| Mikko Arvas | Finnish Red Cross Blood Service / Finnish Hematology Registry and Clinical Biobank, | | Scientific Committee | University of Helsinki & Biobanks |
| Olli Carpén | Helsinki Biobank / Helsinki University and Hospital District of Helsinki and Uusimaa, | | Scientific Committee | University of Helsinki & Biobanks |
| Reetta Hinttala | Northern Finland Biobank Borealis / University of Oulu / Northern Ostrobothnia Hospital | | Scientific Committee | University of Helsinki & Biobanks |
| Johannes Kettunen | Northern Finland Biobank Borealis / University of Oulu / Northern Ostrobothnia Hospital | | Scientific Committee | University of Helsinki & Biobanks |
| Arto Mannermaa | Biobank of Eastern Finland / University of Eastern Finland / Northern Savo Hospital | | Scientific Committee | University of Helsinki & Biobanks |
| Katriina Aalto-Setälä | Faculty of Medicine and Health Technology, Tampere University, Tampere, Finland | | Scientific Committee | University of Helsinki & Biobanks |
| Mika Kähönen | Finnish Clinical Biobank Tampere / University of Tampere / Pirkanmaa Hospital | | Scientific Committee | University of Helsinki & Biobanks |
| Jari Laukkanen | Central Finland Biobank / University of Jyväskylä / Central Finland Health Care District, | | Scientific Committee | University of Helsinki & Biobanks |
| Johanna Mäkelä | FINBB - Finnish biobank cooperative | | Scientific Committee | University of Helsinki & Biobanks |
| Reetta Kälviäinen | Northern Savo Hospital District, Kuopio, Finland | | Clinical Groups | Neurology Group |
| Valtteri Julkunen | Northern Savo Hospital District, Kuopio, Finland | | Clinical Groups | Neurology Group |
| Hilkka Soininen | Northern Savo Hospital District, Kuopio, Finland | | Clinical Groups | Neurology Group |
| Anne Remes | Northern Ostrobothnia Hospital District, Oulu, Finland | | Clinical Groups | Neurology Group |
| Mikko Hiltunen | University of Eastern Finland, Kuopio, Finland | | Clinical Groups | Neurology Group |
| Jukka Peltola | Pirkanmaa Hospital District, Tampere, Finland | | Clinical Groups | Neurology Group |
| Minna Raivio | Hospital District of Helsinki and Uusimaa, Helsinki, Finland | | Clinical Groups | Neurology Group |
| Pentti Tienari | Hospital District of Helsinki and Uusimaa, Helsinki, Finland | | Clinical Groups | Neurology Group |
| Juha Rinne | Hospital District of Southwest Finland, Turku, Finland | | Clinical Groups | Neurology Group |
| Roosa Kallionpää | Hospital District of Southwest Finland, Turku, Finland | | Clinical Groups | Neurology Group |
| Juulia Partanen | Institute for Molecular Medicine Finland, HiLIFE, University of Helsinki, Finland | | Clinical Groups | Neurology Group |
| Adam Ziemann | Abbvie, Chicago, IL, United States | | Clinical Groups | Neurology Group |
| Nizar Smaoui | Abbvie, Chicago, IL, United States | | Clinical Groups | Neurology Group |
| Anne Lehtonen | Abbvie, Chicago, IL, United States | | Clinical Groups | Neurology Group |
| Susan Eaton | Biogen, Cambridge, MA, United States | | Clinical Groups | Neurology Group |
| Heiko Runz | Biogen, Cambridge, MA, United States | | Clinical Groups | Neurology Group |
| Sanni Lahdenperä | Biogen, Cambridge, MA, United States | | Clinical Groups | Neurology Group |
| Shameek Biswas | Bristol Myers Squibb, New York, NY, United States | | Clinical Groups | Neurology Group |
| Natalie Bowers | Genentech, San Francisco, CA, United States | | Clinical Groups | Neurology Group |
| Edmond Teng | Genentech, San Francisco, CA, United States | | Clinical Groups | Neurology Group |

| | | | | |
|---|---|---|---|---|
| Rion Pendergrass | Genentech, San Francisco, CA, United States | | **Clinical Groups** | **Neurology Group** |
| Fanli Xu | GlaxoSmithKline, Brentford, United Kingdom | | **Clinical Groups** | **Neurology Group** |
| David Pulford | GlaxoSmithKline, Stevenage, United Kingdom | | **Clinical Groups** | **Neurology Group** |
| Kirsi Auro | GlaxoSmithKline, Espoo, Finland | | **Clinical Groups** | **Neurology Group** |
| Laura Addis | GlaxoSmithKline, Brentford, United Kingdom | | **Clinical Groups** | **Neurology Group** |
| John Eicher | GlaxoSmithKline, Brentford, United Kingdom | | **Clinical Groups** | **Neurology Group** |
| Qingqin S Li | Janssen Research & Development, LLC, Titusville, NJ 08560, United States | | **Clinical Groups** | **Neurology Group** |
| Karen He | Janssen Research & Development, LLC, Spring House, PA, United States | | **Clinical Groups** | **Neurology Group** |
| Ekaterina Khramtsova | Janssen Research & Development, LLC, Spring House, PA, United States | | **Clinical Groups** | **Neurology Group** |
| Neha Raghavan | Merck, Kenilworth, NJ, United States | | **Clinical Groups** | **Neurology Group** |
| Martti Färkkilä | Hospital District of Helsinki and Uusimaa, Helsinki, Finland | | **Clinical Groups** | **Gastroenterology Group** |
| Jukka Koskela | Hospital District of Helsinki and Uusimaa, Helsinki, Finland | | **Clinical Groups** | **Gastroenterology Group** |
| Sampsa Pikkarainen | Hospital District of Helsinki and Uusimaa, Helsinki, Finland | | **Clinical Groups** | **Gastroenterology Group** |
| Airi Jussila | Pirkanmaa Hospital District, Tampere, Finland | | **Clinical Groups** | **Gastroenterology Group** |
| Katri Kaukinen | Pirkanmaa Hospital District, Tampere, Finland | | **Clinical Groups** | **Gastroenterology Group** |
| Timo Blomster | Northern Ostrobothnia Hospital District, Oulu, Finland | | **Clinical Groups** | **Gastroenterology Group** |
| Mikko Kiviniemi | Northern Savo Hospital District, Kuopio, Finland | | **Clinical Groups** | **Gastroenterology Group** |
| Markku Voutilainen | Hospital District of Southwest Finland, Turku, Finland | | **Clinical Groups** | **Gastroenterology Group** |
| Mark Daly | Institute for Molecular Medicine, Finland (FIMM), HiLIFE, University of Helsinki, | | **Clinical Groups** | **Gastroenterology Group** |
| Jeffrey Waring | Abbvie, Chicago, IL, United States | | **Clinical Groups** | **Gastroenterology Group** |
| Nizar Smaoui | Abbvie, Chicago, IL, United States | | **Clinical Groups** | **Gastroenterology Group** |
| Fedik Rahimov | Abbvie, Chicago, IL, United States | | **Clinical Groups** | **Gastroenterology Group** |
| Anne Lehtonen | Abbvie, Chicago, IL, United States | | **Clinical Groups** | **Gastroenterology Group** |
| Tim Lu | Genentech, San Francisco, CA, United States | | **Clinical Groups** | **Gastroenterology Group** |
| Natalie Bowers | Genentech, San Francisco, CA, United States | | **Clinical Groups** | **Gastroenterology Group** |
| Rion Pendergrass | Genentech, San Francisco, CA, United States | | **Clinical Groups** | **Gastroenterology Group** |
| Linda McCarthy | GlaxoSmithKline, Brentford, United Kingdom | | **Clinical Groups** | **Gastroenterology Group** |
| Amy Hart | Janssen Research & Development, LLC, Spring House, PA, United States | | **Clinical Groups** | **Gastroenterology Group** |
| Meijian Guan | Janssen Research & Development, LLC, Spring House, PA, United States | | **Clinical Groups** | **Gastroenterology Group** |
| Jason Miller | Merck, Kenilworth, NJ, United States | | **Clinical Groups** | **Gastroenterology Group** |
| Kirsi Kalpala | Pfizer, New York, NY, United States | | **Clinical Groups** | **Gastroenterology Group** |
| Melissa Miller | Pfizer, New York, NY, United States | | **Clinical Groups** | **Gastroenterology Group** |
| Xinli Hu | Pfizer, New York, NY, United States | | **Clinical Groups** | **Gastroenterology Group** |
| Kari Eklund | Hospital District of Helsinki and Uusimaa, Helsinki, Finland | | **Clinical Groups** | **Rheumatology Group** |
| Antti Palomäki | Hospital District of Southwest Finland, Turku, Finland | | **Clinical Groups** | **Rheumatology Group** |
| Pia Isomäki | Pirkanmaa Hospital District, Tampere, Finland | | **Clinical Groups** | **Rheumatology Group** |
| Laura Pirilä | Hospital District of Southwest Finland, Turku, Finland | | **Clinical Groups** | **Rheumatology Group** |
| Oili Kaipiainen-Seppäner | Northern Savo Hospital District, Kuopio, Finland | | **Clinical Groups** | **Rheumatology Group** |
| Johanna Huhtakangas | Northern Ostrobothnia Hospital District, Oulu, Finland | | **Clinical Groups** | **Rheumatology Group** |
| Nina Mars | Institute for Molecular Medicine Finland (FIMM), HiLIFE, University of Helsinki, Helsin | | **Clinical Groups** | **Rheumatology Group** |
| Jeffrey Waring | Abbvie, Chicago, IL, United States | | **Clinical Groups** | **Rheumatology Group** |
| Fedik Rahimov | Abbvie, Chicago, IL, United States | | **Clinical Groups** | **Rheumatology Group** |
| Apinya Lertratanakul | Abbvie, Chicago, IL, United States | | **Clinical Groups** | **Rheumatology Group** |
| Nizar Smaoui | Abbvie, Chicago, IL, United States | | **Clinical Groups** | **Rheumatology Group** |
| Anne Lehtonen | Abbvie, Chicago, IL, United States | | **Clinical Groups** | **Rheumatology Group** |
| Coralie Viollet | AstraZeneca, Cambridge, United Kingdom | | **Clinical Groups** | **Rheumatology Group** |
| Marla Hochfeld | Bristol Myers Squibb, New York, NY, United States | | **Clinical Groups** | **Rheumatology Group** |
| Natalie Bowers | Genentech, San Francisco, CA, United States | | **Clinical Groups** | **Rheumatology Group** |
| Rion Pendergrass | Genentech, San Francisco, CA, United States | | **Clinical Groups** | **Rheumatology Group** |
| Jorge Esparza Gordillo | GlaxoSmithKline, Brentford, United Kingdom | | **Clinical Groups** | **Rheumatology Group** |
| Kirsi Auro | GlaxoSmithKline, Espoo, Finland | | **Clinical Groups** | **Rheumatology Group** |
| Dawn Waterworth | Janssen Research & Development, LLC, Spring House, PA, United States | | **Clinical Groups** | **Rheumatology Group** |
| Fabiana Farias | Merck, Kenilworth, NJ, United States | | **Clinical Groups** | **Rheumatology Group** |
| Kirsi Kalpala | Pfizer, New York, NY, United States | | **Clinical Groups** | **Rheumatology Group** |
| Nan Bing | Pfizer, New York, NY, United States | | **Clinical Groups** | **Rheumatology Group** |
| Xinli Hu | Pfizer, New York, NY, United States | | **Clinical Groups** | **Rheumatology Group** |
| Tarja Laitinen | Pirkanmaa Hospital District, Tampere, Finland | | **Clinical Groups** | **Pulmonology Group** |
| Margit Pelkonen | Northern Savo Hospital District, Kuopio, Finland | | **Clinical Groups** | **Pulmonology Group** |
| Paula Kauppi | Hospital District of Helsinki and Uusimaa, Helsinki, Finland | | **Clinical Groups** | **Pulmonology Group** |
| Hannu Kankaanranta | University of Gothenburg, Gothenburg, Sweden/ Seinäjoki Central Hospital, Seinäjoki, | | **Clinical Groups** | **Pulmonology Group** |
| Terttu Harju | Northern Ostrobothnia Hospital District, Oulu, Finland | | **Clinical Groups** | **Pulmonology Group** |
| Riitta Lahesmaa | Hospital District of Southwest Finland, Turku, Finland | | **Clinical Groups** | **Pulmonology Group** |
| Nizar Smaoui | Abbvie, Chicago, IL, United States | | **Clinical Groups** | **Pulmonology Group** |
| Coralie Viollet | AstraZeneca, Cambridge, United Kingdom | | **Clinical Groups** | **Pulmonology Group** |
| Susan Eaton | Biogen, Cambridge, MA, United States | | **Clinical Groups** | **Pulmonology Group** |
| Hubert Chen | Genentech, San Francisco, CA, United States | | **Clinical Groups** | **Pulmonology Group** |
| Rion Pendergrass | Genentech, San Francisco, CA, United States | | **Clinical Groups** | **Pulmonology Group** |
| Natalie Bowers | Genentech, San Francisco, CA, United States | | **Clinical Groups** | **Pulmonology Group** |
| Joanna Betts | GlaxoSmithKline, Brentford, United Kingdom | | **Clinical Groups** | **Pulmonology Group** |
| Kirsi Auro | GlaxoSmithKline, Espoo, Finland | | **Clinical Groups** | **Pulmonology Group** |
| Rajashree Mishra | GlaxoSmithKline, Brentford, United Kingdom | | **Clinical Groups** | **Pulmonology Group** |
| Majd Mouded | Novartis, Basel, Switzerland | | **Clinical Groups** | **Pulmonology Group** |
| Debby Ngo | Novartis, Basel, Switzerland | | **Clinical Groups** | **Pulmonology Group** |
| Teemu Niiranen | Finnish Institute for Health and Welfare (THL), Helsinki, Finland | | **Clinical Groups** | **Cardiometabolic Diseases Group** |
| Felix Vaura | Finnish Institute for Health and Welfare (THL), Helsinki, Finland | | **Clinical Groups** | **Cardiometabolic Diseases Group** |
| Veikko Salomaa | Finnish Institute for Health and Welfare (THL), Helsinki, Finland | | **Clinical Groups** | **Cardiometabolic Diseases Group** |
| Kaj Metsärinne | Hospital District of Southwest Finland, Turku, Finland | | **Clinical Groups** | **Cardiometabolic Diseases Group** |
| Jenni Aittokallio | Hospital District of Southwest Finland, Turku, Finland | | **Clinical Groups** | **Cardiometabolic Diseases Group** |
| Mika Kähönen | Pirkanmaa Hospital District, Tampere, Finland | | **Clinical Groups** | **Cardiometabolic Diseases Group** |
| Jussi Hernesniemi | Pirkanmaa Hospital District, Tampere, Finland | | **Clinical Groups** | **Cardiometabolic Diseases Group** |
| Daniel Gordin | Hospital District of Helsinki and Uusimaa, Helsinki, Finland | | **Clinical Groups** | **Cardiometabolic Diseases Group** |
| Juha Sinisalo | Hospital District of Helsinki and Uusimaa, Helsinki, Finland | | **Clinical Groups** | **Cardiometabolic Diseases Group** |
| Marja-Riitta Taskinen | Hospital District of Helsinki and Uusimaa, Helsinki, Finland | | **Clinical Groups** | **Cardiometabolic Diseases Group** |
| Tiinamaija Tuomi | Hospital District of Helsinki and Uusimaa, Helsinki, Finland | | **Clinical Groups** | **Cardiometabolic Diseases Group** |
| Timo Hiltunen | Hospital District of Helsinki and Uusimaa, Helsinki, Finland | | **Clinical Groups** | **Cardiometabolic Diseases Group** |
| Jari Laukkanen | Central Finland Health Care District, Jyväskylä, Finland | | **Clinical Groups** | **Cardiometabolic Diseases Group** |
| Amanda Elliott | Institute for Molecular Medicine Finland (FIMM), HiLIFE, University of Helsinki, Helsinki, | | **Clinical Groups** | **Cardiometabolic Diseases Group** |
| Mary Pat Reeve | Institute for Molecular Medicine Finland (FIMM), HiLIFE, University of Helsinki, Helsin | | **Clinical Groups** | **Cardiometabolic Diseases Group** |
| Sanni Ruotsalainen | Institute for Molecular Medicine Finland (FIMM), HiLIFE, University of Helsinki, Helsin | | **Clinical Groups** | **Cardiometabolic Diseases Group** |
| Dirk Paul | Astra Zeneca, Cambridge, United Kingdom | | **Clinical Groups** | **Cardiometabolic Diseases Group** |
| Natalie Bowers | Genentech, San Francisco, CA, United States | | **Clinical Groups** | **Cardiometabolic Diseases Group** |
| Rion Pendergrass | Genentech, San Francisco, CA, United States | | **Clinical Groups** | **Cardiometabolic Diseases Group** |
| Audrey Chu | GlaxoSmithKline, Brentford, United Kingdom | | **Clinical Groups** | **Cardiometabolic Diseases Group** |
| Kirsi Auro | GlaxoSmithKline, Espoo, Finland | | **Clinical Groups** | **Cardiometabolic Diseases Group** |
| Dermot Reilly | Janssen Research & Development, LLC, Boston, MA, United States | | **Clinical Groups** | **Cardiometabolic Diseases Group** |
| Mike Mendelson | Novartis, Boston, MA, United States | | **Clinical Groups** | **Cardiometabolic Diseases Group** |
| Jaakko Parkkinen | Pfizer, New York, NY, United States | | **Clinical Groups** | **Cardiometabolic Diseases Group** |
| Melissa Miller | Pfizer, New York, NY, United States | | **Clinical Groups** | **Cardiometabolic Diseases Group** |
| Tuomo Meretoja | Hospital District of Helsinki and Uusimaa, Helsinki, Finland | | **Clinical Groups** | **Oncology Group** |
| Heikki Joensuu | Hospital District of Helsinki and Uusimaa, Helsinki, Finland | | **Clinical Groups** | **Oncology Group** |
| Olli Carpén | Hospital District of Helsinki and Uusimaa, Helsinki, Finland | | **Clinical Groups** | **Oncology Group** |
| Johanna Mattson | Hospital District of Helsinki and Uusimaa, Helsinki, Finland | | **Clinical Groups** | **Oncology Group** |
| Eveliina Salminen | Hospital District of Helsinki and Uusimaa, Helsinki, Finland | | **Clinical Groups** | **Oncology Group** |

| | | | | |
|---|---|---|---|---|
| Annika Auranen | Pirkanmaa Hospital District , Tampere, Finland | | Clinical Groups | **Oncology Group** |
| Peeter Karihtala | Northern Ostrobothnia Hospital District, Oulu, Finland | | Clinical Groups | **Oncology Group** |
| Päivi Auvinen | Northern Savo Hospital District, Kuopio, Finland | | Clinical Groups | **Oncology Group** |
| Klaus Elenius | Hospital District of Southwest Finland, Turku, Finland | | Clinical Groups | **Oncology Group** |
| Johanna Schleutker | Hospital District of Southwest Finland, Turku, Finland | | Clinical Groups | **Oncology Group** |
| Esa Pitkänen | Institute for Molecular Medicine Finland (FIMM), HiLIFE, University of Helsinki, Helsin | | Clinical Groups | **Oncology Group** |
| Nina Mars | Institute for Molecular Medicine Finland (FIMM), HiLIFE, University of Helsinki, Helsin | | Clinical Groups | **Oncology Group** |
| Mark Daly | Institute for Molecular Medicine Finland (FIMM), HiLIFE, University of Helsinki, Helsinki, | | Clinical Groups | **Oncology Group** |
| Relja Popovic | Abbvie, Chicago, IL, United States | | Clinical Groups | **Oncology Group** |
| Jeffrey Waring | Abbvie, Chicago, IL, United States | | Clinical Groups | **Oncology Group** |
| Bridget Riley-Gillis | Abbvie, Chicago, IL, United States | | Clinical Groups | **Oncology Group** |
| Anne Lehtonen | Abbvie, Chicago, IL, United States | | Clinical Groups | **Oncology Group** |
| Margarete Fabre | AstraZeneca, Cambridge, United Kingdom | | Clinical Groups | **Oncology Group** |
| Jennifer Schutzman | Genentech, San Francisco, CA, United States | | Clinical Groups | **Oncology Group** |
| Natalie Bowers | Genentech, San Francisco, CA, United States | | Clinical Groups | **Oncology Group** |
| Rion Pendergrass | Genentech, San Francisco, CA, United States | | Clinical Groups | **Oncology Group** |
| Diptee Kulkarni | GlaxoSmithKline, Brentford, United Kingdom | | Clinical Groups | **Oncology Group** |
| Kirsi Auro | GlaxoSmithKline, Espoo, Finland | | Clinical Groups | **Oncology Group** |
| Alessandro Porello | Janssen Research & Development, LLC, Spring House, PA, United States | | Clinical Groups | **Oncology Group** |
| Andrey Loboda | Merck, Kenilworth, NJ, United States | | Clinical Groups | **Oncology Group** |
| Heli Lehtonen | Pfizer, New York, NY, United States | | Clinical Groups | **Oncology Group** |
| Stefan McDonough | Pfizer, New York, NY, United States | | Clinical Groups | **Oncology Group** |
| Sauli Vuoti | Janssen-Cilag Oy, Espoo, Finland | | Clinical Groups | **Oncology Group** |
| Kai Kaarniranta | Northern Savo Hospital District, Kuopio, Finland; Department of Molecular Genetics, | | Clinical Groups | **Opthalmology Group** |
| Joni A Turunen | Helsinki University Hospital and University of Helsinki, Helsinki, Finland; Eye Genetics | | Clinical Groups | **Opthalmology Group** |
| Terhi Ollila | Hospital District of Helsinki and Uusimaa, Helsinki, Finland | | Clinical Groups | **Opthalmology Group** |
| Hannu Uusitalo | Pirkanmaa Hospital District, Tampere, Finland | | Clinical Groups | **Opthalmology Group** |
| Juha Karjalainen | Institute for Molecular Medicine Finland (FIMM), HiLIFE, University of Helsinki, Helsin | | Clinical Groups | **Opthalmology Group** |
| Esa Pitkänen | Institute for Molecular Medicine Finland (FIMM), HiLIFE, University of Helsinki, Helsin | | Clinical Groups | **Opthalmology Group** |
| Mengzhen Liu | Abbvie, Chicago, IL, United States | | Clinical Groups | **Opthalmology Group** |
| Heiko Runz | Biogen, Cambridge, MA, United States | | Clinical Groups | **Opthalmology Group** |
| Stephanie Loomis | Biogen, Cambridge, MA, United States | | Clinical Groups | **Opthalmology Group** |
| Erich Strauss | Genentech, San Francisco, CA, United States | | Clinical Groups | **Opthalmology Group** |
| Natalie Bowers | Genentech, San Francisco, CA, United States | | Clinical Groups | **Opthalmology Group** |
| Hao Chen | Genentech, San Francisco, CA, United States | | Clinical Groups | **Opthalmology Group** |
| Rion Pendergrass | Genentech, San Francisco, CA, United States | | Clinical Groups | **Opthalmology Group** |
| Kaisa Tasanen | Northern Ostrobothnia Hospital District, Oulu, Finland | | Clinical Groups | **Dermatology Group** |
| Laura Huilaja | Northern Ostrobothnia Hospital District, Oulu, Finland | | Clinical Groups | **Dermatology Group** |
| Katariina Hannula-Joupp | Hospital District of Helsinki and Uusimaa, Helsinki, Finland | | Clinical Groups | **Dermatology Group** |
| Teea Salmi | Pirkanmaa Hospital District, Tampere, Finland | | Clinical Groups | **Dermatology Group** |
| Sirkku Peltonen | Hospital District of Southwest Finland, Turku, Finland | | Clinical Groups | **Dermatology Group** |
| Leena Koulu | Hospital District of Southwest Finland, Turku, Finland | | Clinical Groups | **Dermatology Group** |
| Nizar Smaoui | Abbvie, Chicago, IL, United States | | Clinical Groups | **Dermatology Group** |
| Fedik Rahimov | Abbvie, Chicago, IL, United States | | Clinical Groups | **Dermatology Group** |
| Anne Lehtonen | Abbvie, Chicago, IL, United States | | Clinical Groups | **Dermatology Group** |
| David Choy | Genentech, San Francisco, CA, United States | | Clinical Groups | **Dermatology Group** |
| Rion Pendergrass | Genentech, San Francisco, CA, United States | | Clinical Groups | **Dermatology Group** |
| Dawn Waterworth | Janssen Research & Development, LLC, Spring House, PA, United States | | Clinical Groups | **Dermatology Group** |
| Kirsi Kalpala | Pfizer, New York, NY, United States | | Clinical Groups | **Dermatology Group** |
| Ying Wu | Pfizer, New York, NY, United States | | Clinical Groups | **Dermatology Group** |
| Pirkko Pussinen | Hospital District of Helsinki and Uusimaa, Helsinki, Finland | | Clinical Groups | **Odontology Group** |
| Aino Salminen | Hospital District of Helsinki and Uusimaa, Helsinki, Finland | | Clinical Groups | **Odontology Group** |
| Tuula Salo | Hospital District of Helsinki and Uusimaa, Helsinki, Finland | | Clinical Groups | **Odontology Group** |
| David Rice | Hospital District of Helsinki and Uusimaa, Helsinki, Finland | | Clinical Groups | **Odontology Group** |
| Pekka Nieminen | Hospital District of Helsinki and Uusimaa, Helsinki, Finland | | Clinical Groups | **Odontology Group** |
| Ulla Palotie | Hospital District of Helsinki and Uusimaa, Helsinki, Finland | | Clinical Groups | **Odontology Group** |
| Maria Siponen | Northern Savo Hospital District, Kuopio, Finland | | Clinical Groups | **Odontology Group** |
| Liisa Suominen | Northern Savo Hospital District, Kuopio, Finland | | Clinical Groups | **Odontology Group** |
| Päivi Mäntylä | Northern Savo Hospital District, Kuopio, Finland | | Clinical Groups | **Odontology Group** |
| Ulvi Gursoy | Hospital District of Southwest Finland, Turku, Finland | | Clinical Groups | **Odontology Group** |
| Vuokko Anttonen | Northern Ostrobothnia Hospital District, Oulu, Finland | | Clinical Groups | **Odontology Group** |
| Kirsi Sipilä | Research Unit of Oral Health Sciences Faculty of Medicine, University of Oulu, Oulu, | | Clinical Groups | **Odontology Group** |
| Rion Pendergrass | Genentech, San Francisco, CA, United States | | Clinical Groups | **Odontology Group** |
| Hannele Laivuori | Institute for Molecular Medicine Finland (FIMM), HiLIFE, University of Helsinki, Helsin | | Clinical Groups | **Women's Health and Reproduction Group** |
| Venla Kurra | Pirkanmaa Hospital District, Tampere, Finland | | Clinical Groups | **Women's Health and Reproduction Group** |
| Laura Kotaniemi-Taloner | Pirkanmaa Hospital District, Tampere, Finland | | Clinical Groups | **Women's Health and Reproduction Group** |
| Oskari Heikinheimo | Hospital District of Helsinki and Uusimaa, Helsinki, Finland | | Clinical Groups | **Women's Health and Reproduction Group** |
| Ilkka Kalliala | Hospital District of Helsinki and Uusimaa, Helsinki, Finland | | Clinical Groups | **Women's Health and Reproduction Group** |
| Lauri Aaltonen | Hospital District of Helsinki and Uusimaa, Helsinki, Finland | | Clinical Groups | **Women's Health and Reproduction Group** |
| Varpu Jokimaa | Hospital District of Southwest Finland, Turku, Finland | | Clinical Groups | **Women's Health and Reproduction Group** |
| Johannes Kettunen | Northern Ostrobothnia Hospital District, Oulu, Finland | | Clinical Groups | **Women's Health and Reproduction Group** |
| Marja Vääräsmäki | Northern Ostrobothnia Hospital District, Oulu, Finland | | Clinical Groups | **Women's Health and Reproduction Group** |
| Outi Uimari | Northern Ostrobothnia Hospital District, Oulu, Finland | | Clinical Groups | **Women's Health and Reproduction Group** |
| Laure Morin-Papunen | Northern Ostrobothnia Hospital District, Oulu, Finland | | Clinical Groups | **Women's Health and Reproduction Group** |
| Maarit Niinimäki | Northern Ostrobothnia Hospital District, Oulu, Finland | | Clinical Groups | **Women's Health and Reproduction Group** |
| Terhi Piltonen | Northern Ostrobothnia Hospital District, Oulu, Finland | | Clinical Groups | **Women's Health and Reproduction Group** |
| Katja Kivinen | Institute for Molecular Medicine Finland (FIMM), HiLIFE, University of Helsinki, Helsin | | Clinical Groups | **Women's Health and Reproduction Group** |
| Elisabeth Widen | Institute for Molecular Medicine Finland (FIMM), HiLIFE, University of Helsinki, Helsin | | Clinical Groups | **Women's Health and Reproduction Group** |
| Taru Tukiainen | Institute for Molecular Medicine Finland (FIMM), HiLIFE, University of Helsinki, Helsin | | Clinical Groups | **Women's Health and Reproduction Group** |
| Mary Pat Reeve | Institute for Molecular Medicine Finland (FIMM), HiLIFE, University of Helsinki, Helsin | | Clinical Groups | **Women's Health and Reproduction Group** |
| Mark Daly | Institute for Molecular Medicine Finland (FIMM), HiLIFE, University of Helsinki, Helsinki, | | Clinical Groups | **Women's Health and Reproduction Group** |
| Niko Välimäki | University of Helsinki, Helsinki, Finland | | Clinical Groups | **Women's Health and Reproduction Group** |
| Eija Laakkonen | University of Jyväskylä, Jyväskylä, Finland | | Clinical Groups | **Women's Health and Reproduction Group** |
| Jaakko Tyrmi | University of Oulu, Oulu, Finland / University of Tampere, Tampere, Finland | | Clinical Groups | **Women's Health and Reproduction Group** |
| Heidi Silven | University of Oulu, Oulu, Finland | | Clinical Groups | **Women's Health and Reproduction Group** |
| Eeva Sliz | University of Oulu, Oulu, Finland | | Clinical Groups | **Women's Health and Reproduction Group** |
| Riikka Arffman | University of Oulu, Oulu, Finland | | Clinical Groups | **Women's Health and Reproduction Group** |
| Susanna Savukoski | University of Oulu, Oulu, Finland | | Clinical Groups | **Women's Health and Reproduction Group** |
| Triin Laisk | Estonian biobank, Tartu, Estonia | | Clinical Groups | **Women's Health and Reproduction Group** |
| Natalia Pujol | Estonian biobank, Tartu, Estonia | | Clinical Groups | **Women's Health and Reproduction Group** |
| Mengzhen Liu | Abbvie, Chicago, IL, United States | | Clinical Groups | **Women's Health and Reproduction Group** |
| Bridget Riley-Gillis | Abbvie, Chicago, IL, United States | | Clinical Groups | **Women's Health and Reproduction Group** |
| Rion Pendergrass | Genentech, San Francisco, CA, United States | | Clinical Groups | **Women's Health and Reproduction Group** |
| Janet Kumar | GlaxoSmithKline, Collegeville, PA, United States | | Clinical Groups | **Women's Health and Reproduction Group** |
| Kirsi Auro | GlaxoSmithKline, Espoo, Finland | | Clinical Groups | **Women's Health and Reproduction Group** |
| Iiris Hovatta | University of Helsinki, Finland | | Clinical Groups | **Depression group** |
| Chia-Yen Chen | Biogen, Cambridge, MA, United States | | Clinical Groups | **Depression group** |
| Erkki Isometsä | Hospital District of Helsinki and Uusimaa, Helsinki, Finland | | Clinical Groups | **Depression group** |
| Hanna Ollila | Institute for Molecular Medicine Finland (FIMM), HiLIFE, University of Helsinki, Helsin | | Clinical Groups | **Depression group** |
| Jaana Suvisaari | Finnish Institute for Health and Welfare (THL), Helsinki, Finland | | Clinical Groups | **Depression group** |
| Antti Mäkitie | Department of Otorhinolaryngology - Head and Neck Surgery, University of Helsinki | | Clinical Groups | **ENT (ear, nose and throath) Group** |
| Argyro Bizaki-Vallaskang | Pirkanmaa Hospital District, Tampere, Finland | | Clinical Groups | **ENT (ear, nose and throath) Group** |
| Sanna Toppila-Salmi | University of Eastern Finland and Kuopio University Hospital, Department of | | Clinical Groups | **ENT (ear, nose and throath) Group** |

| | | | | |
|---|---|---|---|---|
| Tytti Willberg | Hospital District of Southwest Finland, Turku, Finland | | Clinical Groups | **ENT (ear, nose and throath) Group** |
| Elmo Saarentaus | Institute for Molecular Medicine Finland (FIMM), HiLIFE, University of Helsinki, Helsin | | Clinical Groups | **ENT (ear, nose and throath) Group** |
| Antti Aarnisalo | Hospital District of Helsinki and Uusimaa, Helsinki, Finland | | Clinical Groups | **ENT (ear, nose and throath) Group** |
| Eveliina Salminen | Hospital District of Helsinki and Uusimaa, Helsinki, Finland | | Clinical Groups | **ENT (ear, nose and throath) Group** |
| Elisa Rahikkala | Northern Ostrobothnia Hospital District, Oulu, Finland | | Clinical Groups | **ENT (ear, nose and throath) Group** |
| Johannes Kettunen | Northern Ostrobothnia Hospital District, Oulu, Finland | | Clinical Groups | **ENT (ear, nose and throath) Group** |
| Kristiina Aittomäki | Department of Medical Genetics, Helsinki University Central Hospital, Helsinki, Finland | | Clinical Groups | **POI (premature ovarian failure) Group** |
| Fredrik Åberg | Transplantation and Liver Surgery Clinic, Helsinki University Hospital, Helsinki | | Clinical Groups | **LiverScore Group** |
| Mitja Kurki | Institute for Molecular Medicine Finland (FIMM), HiLIFE, University of Helsinki, Helsinki, | | FinnGen Analysis working gr | FinnGen Analysis working group |
| Samuli Ripatti | Institute for Molecular Medicine Finland (FIMM), HiLIFE, University of Helsinki, Helsin | | FinnGen Analysis working gr | FinnGen Analysis working group |
| Mark Daly | Institute for Molecular Medicine, Finland (FIMM), HiLIFE, University of Helsinki, | | FinnGen Analysis working gr | FinnGen Analysis working group |
| Juha Karjalainen | Institute for Molecular Medicine Finland (FIMM), HiLIFE, University of Helsinki, Helsin | | FinnGen Analysis working gr | FinnGen Analysis working group |
| Aki Havulinna | Institute for Molecular Medicine Finland (FIMM), HiLIFE, University of Helsinki, Helsinki, | | FinnGen Analysis working gr | FinnGen Analysis working group |
| Juha Mehtonen | Institute for Molecular Medicine Finland (FIMM), HiLIFE, University of Helsinki, Helsin | | FinnGen Analysis working gr | FinnGen Analysis working group |
| Priit Palta | Institute for Molecular Medicine Finland (FIMM), HiLIFE, University of Helsinki, Helsin | | FinnGen Analysis working gr | FinnGen Analysis working group |
| Shabbeer Hassan | Institute for Molecular Medicine Finland (FIMM), HiLIFE, University of Helsinki, Helsin | | FinnGen Analysis working gr | FinnGen Analysis working group |
| Pietro Della Briotta Parol | Institute for Molecular Medicine Finland (FIMM), HiLIFE, University of Helsinki, Helsin | | FinnGen Analysis working gr | FinnGen Analysis working group |
| Wei Zhou | Broad Institute, Cambridge, MA, United States | | FinnGen Analysis working gr | FinnGen Analysis working group |
| Mutaamba Maasha | Broad Institute, Cambridge, MA, United States | | FinnGen Analysis working gr | FinnGen Analysis working group |
| Shabbeer Hassan | Institute for Molecular Medicine Finland (FIMM), HiLIFE, University of Helsinki, Helsin | | FinnGen Analysis working gr | FinnGen Analysis working group |
| Susanna Lemmelä | Institute for Molecular Medicine Finland (FIMM), HiLIFE, University of Helsinki, Helsin | | FinnGen Analysis working gr | FinnGen Analysis working group |
| Manuel Rivas | University of Stanford, Stanford, CA, United States | | FinnGen Analysis working gr | FinnGen Analysis working group |
| Aarno Palotie | Institute for Molecular Medicine Finland (FIMM), HiLIFE, University of Helsinki, Helsin | | FinnGen Analysis working gr | FinnGen Analysis working group |
| Aoxing Liu | Institute for Molecular Medicine Finland (FIMM), HiLIFE, University of Helsinki, Helsin | | FinnGen Analysis working gr | FinnGen Analysis working group |
| Arto Lehisto | Institute for Molecular Medicine Finland (FIMM), HiLIFE, University of Helsinki, Helsin | | FinnGen Analysis working gr | FinnGen Analysis working group |
| Andrea Ganna | Institute for Molecular Medicine Finland (FIMM), HiLIFE, University of Helsinki, Helsin | | FinnGen Analysis working gr | FinnGen Analysis working group |
| Vincent Llorens | Institute for Molecular Medicine Finland (FIMM), HiLIFE, University of Helsinki, Helsin | | FinnGen Analysis working gr | FinnGen Analysis working group |
| Hannele Laivuori | Institute for Molecular Medicine Finland (FIMM), HiLIFE, University of Helsinki, Helsin | | FinnGen Analysis working gr | FinnGen Analysis working group |
| Taru Tukiainen | Institute for Molecular Medicine Finland (FIMM), HiLIFE, University of Helsinki, Helsin | | FinnGen Analysis working gr | FinnGen Analysis working group |
| Mary Pat Reeve | Institute for Molecular Medicine Finland (FIMM), HiLIFE, University of Helsinki, Helsin | | FinnGen Analysis working gr | FinnGen Analysis working group |
| Henrike Heyne | Institute for Molecular Medicine Finland (FIMM), HiLIFE, University of Helsinki, Helsin | | FinnGen Analysis working gr | FinnGen Analysis working group |
| Nina Mars | Institute for Molecular Medicine Finland (FIMM), HiLIFE, University of Helsinki, Helsin | | FinnGen Analysis working gr | FinnGen Analysis working group |
| Joel Rämö | Institute for Molecular Medicine Finland (FIMM), HiLIFE, University of Helsinki, Helsin | | FinnGen Analysis working gr | FinnGen Analysis working group |
| Elmo Saarentaus | Institute for Molecular Medicine Finland (FIMM), HiLIFE, University of Helsinki, Helsin | | FinnGen Analysis working gr | FinnGen Analysis working group |
| Hanna Ollila | Institute for Molecular Medicine Finland (FIMM), HiLIFE, University of Helsinki, Helsin | | FinnGen Analysis working gr | FinnGen Analysis working group |
| Rodos Rodosthenous | Institute for Molecular Medicine Finland (FIMM), HiLIFE, University of Helsinki, Helsin | | FinnGen Analysis working gr | FinnGen Analysis working group |
| Satu Strausz | Institute for Molecular Medicine Finland (FIMM), HiLIFE, University of Helsinki, Helsin | | FinnGen Analysis working gr | FinnGen Analysis working group |
| Tuula Palotie | University of Helsinki and Hospital District of Helsinki and Uusimaa, Helsinki, Finland | | FinnGen Analysis working gr | FinnGen Analysis working group |
| Kimmo Palin | University of Helsinki, Helsinki, Finland | | FinnGen Analysis working gr | FinnGen Analysis working group |
| Javier Garcia-Tabuenca | University of Tampere, Tampere, Finland | | FinnGen Analysis working gr | FinnGen Analysis working group |
| Harri Siirtola | University of Tampere, Tampere, Finland | | FinnGen Analysis working gr | FinnGen Analysis working group |
| Tuomo Kiiskinen | Institute for Molecular Medicine Finland (FIMM), HiLIFE, University of Helsinki, Helsin | | FinnGen Analysis working gr | FinnGen Analysis working group |
| Jiwoo Lee | Institute for Molecular Medicine Finland (FIMM), HiLIFE, University of Helsinki, Helsinki, | | FinnGen Analysis working gr | FinnGen Analysis working group |
| Kristin Tsuo | Institute for Molecular Medicine Finland (FIMM), HiLIFE, University of Helsinki, Helsinki, | | FinnGen Analysis working gr | FinnGen Analysis working group |
| Amanda Elliott | Institute for Molecular Medicine Finland (FIMM), HiLIFE, University of Helsinki, Helsinki, | | FinnGen Analysis working gr | FinnGen Analysis working group |
| Kati Kristiansson | THL Biobank / Finnish Institute for Health and Welfare (THL), Helsinki, Finland | | FinnGen Analysis working gr | FinnGen Analysis working group |
| Mikko Arvas | Finnish Red Cross Blood Service / Finnish Hematology Registry and Clinical Biobank, | | FinnGen Analysis working gr | FinnGen Analysis working group |
| Kati Hyvärinen | Finnish Red Cross Blood Service, Helsinki, Finland | | FinnGen Analysis working gr | FinnGen Analysis working group |
| Jarmo Ritari | Finnish Red Cross Blood Service, Helsinki, Finland | | FinnGen Analysis working gr | FinnGen Analysis working group |
| Olli Carpén | Helsinki Biobank / Helsinki University and Hospital District of Helsinki and Uusimaa, | | FinnGen Analysis working gr | FinnGen Analysis working group |
| Johannes Kettunen | Northern Finland Biobank Borealis / University of Oulu / Northern Ostrobothnia Hospital | | FinnGen Analysis working gr | FinnGen Analysis working group |
| Katri Pylkäs | University of Oulu, Oulu, Finland | | FinnGen Analysis working gr | FinnGen Analysis working group |
| Eeva Sliz | University of Oulu, Oulu, Finland | | FinnGen Analysis working gr | FinnGen Analysis working group |
| Minna Karjalainen | University of Oulu, Oulu, Finland | | FinnGen Analysis working gr | FinnGen Analysis working group |
| Tuomo Mantere | Northern Finland Biobank Borealis / University of Oulu / Northern Ostrobothnia Hospital | | FinnGen Analysis working gr | FinnGen Analysis working group |
| Eeva Kangasniemi | Finnish Clinical Biobank Tampere / University of Tampere / Pirkanmaa Hospital | | FinnGen Analysis working gr | FinnGen Analysis working group |
| Sami Heikkinen | University of Eastern Finland, Kuopio, Finland | | FinnGen Analysis working gr | FinnGen Analysis working group |
| Arto Mannermaa | Biobank of Eastern Finland / University of Eastern Finland / Northern Savo Hospital | | FinnGen Analysis working gr | FinnGen Analysis working group |
| Eija Laakkonen | University of Jyväskylä, Jyväskylä, Finland | | FinnGen Analysis working gr | FinnGen Analysis working group |
| Nina Pitkänen | Auria Biobank / University of Turku / Hospital District of Southwest Finland, Turku, | | FinnGen Analysis working gr | FinnGen Analysis working group |
| Samuel Lessard | Translational Sciences, Sanofi R&D, Framingham, MA, USA | | FinnGen Analysis working gr | FinnGen Analysis working group |
| Clément Chatelain | Translational Sciences, Sanofi R&D, Framingham, MA, USA | | FinnGen Analysis working gr | FinnGen Analysis working group |
| Lila Kallio | Auria Biobank / University of Turku / Hospital District of Southwest Finland, Turku, | | Biobank directors | Biobank directors |
| Tiina Wahlfors | THL Biobank / Finnish Institute for Health and Welfare (THL), Helsinki, Finland | | Biobank directors | Biobank directors |
| Jukka Partanen | Finnish Red Cross Blood Service / Finnish Hematology Registry and Clinical Biobank, | | Biobank directors | Biobank directors |
| Eero Punkka | Helsinki Biobank / Helsinki University and Hospital District of Helsinki and Uusimaa, | | Biobank directors | Biobank directors |
| Raisa Serpi | Northern Finland Biobank Borealis / University of Oulu / Northern Ostrobothnia Hospital | | Biobank directors | Biobank directors |
| Sanna Siltanen | Finnish Clinical Biobank Tampere / University of Tampere / Pirkanmaa Hospital | | Biobank directors | Biobank directors |
| Veli-Matti Kosma | Biobank of Eastern Finland / University of Eastern Finland / Northern Savo Hospital | | Biobank directors | Biobank directors |
| Teijo Kuopio | Central Finland Biobank / University of Jyväskylä / Central Finland Health Care District, | | Biobank directors | Biobank directors |
| Anu Jalanko | Institute for Molecular Medicine Finland (FIMM), HiLIFE, University of Helsinki, Helsin | | FinnGen Teams | **Administration** |
| Huei-Yi Shen | Institute for Molecular Medicine Finland (FIMM), HiLIFE, University of Helsinki, Helsin | | FinnGen Teams | **Administration** |
| Risto Kajanne | Institute for Molecular Medicine Finland (FIMM), HiLIFE, University of Helsinki, Helsin | | FinnGen Teams | **Administration** |
| Mervi Aavikko | Institute for Molecular Medicine Finland (FIMM), HiLIFE, University of Helsinki, Helsin | | FinnGen Teams | **Administration** |
| Helen Cooper | Institute for Molecular Medicine Finland (FIMM), HiLIFE, University of Helsinki, Helsin | | FinnGen Teams | **Administration** |
| Denise Öller | Institute for Molecular Medicine Finland (FIMM), HiLIFE, University of Helsinki, Helsin | | FinnGen Teams | **Administration** |
| Rasko Leinonen | Institute for Molecular Medicine Finland (FIMM), HiLIFE, University of Helsinki, Helsin | | FinnGen Teams | **Administration** |
| Henna Palin | Finnish Clinical Biobank Tampere / University of Tampere / Pirkanmaa Hospital | | FinnGen Teams | **Administration** |
| Malla-Maria Linna | Helsinki Biobank / Helsinki University and Hospital District of Helsinki and Uusimaa, | | FinnGen Teams | **Administration** |
| Mitja Kurki | Institute for Molecular Medicine Finland (FIMM), HiLIFE, University of Helsinki, Helsinki, | | FinnGen Teams | **Analysis** |
| Juha Karjalainen | Institute for Molecular Medicine Finland (FIMM), HiLIFE, University of Helsinki, Helsin | | FinnGen Teams | **Analysis** |
| Pietro Della Briotta Parol | Institute for Molecular Medicine Finland (FIMM), HiLIFE, University of Helsinki, Helsin | | FinnGen Teams | **Analysis** |
| Arto Lehisto | Institute for Molecular Medicine Finland (FIMM), HiLIFE, University of Helsinki, Helsin | | FinnGen Teams | **Analysis** |
| Juha Mehtonen | Institute for Molecular Medicine Finland (FIMM), HiLIFE, University of Helsinki, Helsin | | FinnGen Teams | **Analysis** |
| Wei Zhou | Broad Institute, Cambridge, MA, United States | | FinnGen Teams | **Analysis** |
| Masahiro Kanai | Broad Institute, Cambridge, MA, United States | | FinnGen Teams | **Analysis** |
| Mutaamba Maasha | Broad Institute, Cambridge, MA, United States | | FinnGen Teams | **Analysis** |
| Zhili Zheng | Broad Institute, Cambridge, MA, United States | | FinnGen Teams | **Analysis** |
| Hannele Laivuori | Institute for Molecular Medicine Finland (FIMM), HiLIFE, University of Helsinki, Helsin | | FinnGen Teams | **Clinical Endpoint Development** |
| Aki Havulinna | Institute for Molecular Medicine Finland (FIMM), HiLIFE, University of Helsinki, Helsinki, | | FinnGen Teams | **Clinical Endpoint Development** |
| Susanna Lemmelä | Institute for Molecular Medicine Finland (FIMM), HiLIFE, University of Helsinki, Helsin | | FinnGen Teams | **Clinical Endpoint Development** |
| Tuomo Kiiskinen | Institute for Molecular Medicine Finland (FIMM), HiLIFE, University of Helsinki, Helsin | | FinnGen Teams | **Clinical Endpoint Development** |
| L. Elisa Lahtela | Institute for Molecular Medicine Finland (FIMM), HiLIFE, University of Helsinki, Helsin | | FinnGen Teams | **Clinical Endpoint Development** |
| Mari Kaunisto | Institute for Molecular Medicine Finland (FIMM), HiLIFE, University of Helsinki, Helsin | | FinnGen Teams | **Communication** |
| Elina Kilpeläinen | Institute for Molecular Medicine Finland (FIMM), HiLIFE, University of Helsinki, Helsin | | FinnGen Teams | **E-Science** |
| Timo P. Sipilä | Institute for Molecular Medicine Finland (FIMM), HiLIFE, University of Helsinki, Helsin | | FinnGen Teams | **E-Science** |
| Oluwaseun Alexander Da | Institute for Molecular Medicine Finland (FIMM), HiLIFE, University of Helsinki, Helsin | | FinnGen Teams | **E-Science** |
| Awaisa Ghazal | Institute for Molecular Medicine Finland (FIMM), HiLIFE, University of Helsinki, Helsin | | FinnGen Teams | **E-Science** |
| Anastasia Kytölä | Institute for Molecular Medicine Finland (FIMM), HiLIFE, University of Helsinki, Helsin | | FinnGen Teams | **E-Science** |
| Rigbe Weldatsadik | Institute for Molecular Medicine Finland (FIMM), HiLIFE, University of Helsinki, Helsin | | FinnGen Teams | **E-Science** |
| Sanni Ruotsalainen | Institute for Molecular Medicine Finland (FIMM), HiLIFE, University of Helsinki, Helsin | | FinnGen Teams | **E-Science** |
| Kati Donner | Institute for Molecular Medicine Finland (FIMM), HiLIFE, University of Helsinki, Helsin | | FinnGen Teams | **Genotyping** |
| Timo P. Sipilä | Institute for Molecular Medicine Finland (FIMM), HiLIFE, University of Helsinki, Helsin | | FinnGen Teams | **Genotyping** |

| | | | | |
|---|---|---|---|---|
| Anu Loukola | Helsinki Biobank / Helsinki University and Hospital District of Helsinki and Uusimaa, | | **FinnGen Teams** | **Sample Collection Coordination** |
| Päivi Laiho | THL Biobank / Finnish Institute for Health and Welfare (THL), Helsinki, Finland | | **FinnGen Teams** | **Sample Logistics** |
| Tuuli Sistonen | THL Biobank / Finnish Institute for Health and Welfare (THL), Helsinki, Finland | | **FinnGen Teams** | **Sample Logistics** |
| Essi Kaiharju | THL Biobank / Finnish Institute for Health and Welfare (THL), Helsinki, Finland | | **FinnGen Teams** | **Sample Logistics** |
| Markku Laukkanen | THL Biobank / Finnish Institute for Health and Welfare (THL), Helsinki, Finland | | **FinnGen Teams** | **Sample Logistics** |
| Elina Järvensivu | THL Biobank / Finnish Institute for Health and Welfare (THL), Helsinki, Finland | | **FinnGen Teams** | **Sample Logistics** |
| Sini Lähteenmäki | THL Biobank / Finnish Institute for Health and Welfare (THL), Helsinki, Finland | | **FinnGen Teams** | **Sample Logistics** |
| Lotta Männikkö | THL Biobank / Finnish Institute for Health and Welfare (THL), Helsinki, Finland | | **FinnGen Teams** | **Sample Logistics** |
| Regis Wong | THL Biobank / Finnish Institute for Health and Welfare (THL), Helsinki, Finland | | **FinnGen Teams** | **Sample Logistics** |
| Auli Toivola | THL Biobank / Finnish Institute for Health and Welfare (THL), Helsinki, Finland | | **FinnGen Teams** | **Sample Logistics** |
| Minna Brunfeldt | THL Biobank / Finnish Institute for Health and Welfare (THL), Helsinki, Finland | | **FinnGen Teams** | **Registry Data Operations** |
| Hannele Mattsson | THL Biobank / Finnish Institute for Health and Welfare (THL), Helsinki, Finland | | **FinnGen Teams** | **Registry Data Operations** |
| Kati Kristiansson | THL Biobank / Finnish Institute for Health and Welfare (THL), Helsinki, Finland | | **FinnGen Teams** | **Registry Data Operations** |
| Susanna Lemmelä | Institute for Molecular Medicine Finland (FIMM), HiLIFE, University of Helsinki, Helsinl | | **FinnGen Teams** | **Registry Data Operations** |
| Sami Koskelainen | THL Biobank / Finnish Institute for Health and Welfare (THL), Helsinki, Finland | | **FinnGen Teams** | **Registry Data Operations** |
| Tero Hiekkalinna | THL Biobank / Finnish Institute for Health and Welfare (THL), Helsinki, Finland | | **FinnGen Teams** | **Registry Data Operations** |
| Teemu Paajanen | THL Biobank / Finnish Institute for Health and Welfare (THL), Helsinki, Finland | | **FinnGen Teams** | **Registry Data Operations** |
| Priit Palta | Institute for Molecular Medicine Finland (FIMM), HiLIFE, University of Helsinki, Helsinl | | **FinnGen Teams** | **Sequencing Informatics** |
| Shuang Luo | Institute for Molecular Medicine Finland (FIMM), HiLIFE, University of Helsinki, Helsinl | | **FinnGen Teams** | **Sequencing Informatics** |
| Tarja Laitinen | Pirkanmaa Hospital District, Tampere, Finland | | **FinnGen Teams** | **Trajectory** |
| Mary Pat Reeve | Institute for Molecular Medicine Finland (FIMM), HiLIFE, University of Helsinki, Helsinl | | **FinnGen Teams** | **Trajectory** |
| Shanmukha Sampath Pa | Institute for Molecular Medicine Finland (FIMM), HiLIFE, University of Helsinki, Helsinl | | **FinnGen Teams** | **Trajectory** |
| Marianna Niemi | University of Tampere, Tampere, Finland | | **FinnGen Teams** | **Trajectory** |
| Harri Siirtola | University of Tampere, Tampere, Finland | | **FinnGen Teams** | **Trajectory** |
| Javier Gracia-Tabuenca | University of Tampere, Tampere, Finland | | **FinnGen Teams** | **Trajectory** |
| Mika Helminen | University of Tampere, Tampere, Finland | | **FinnGen Teams** | **Trajectory** |
| Tiina Luukkaala | University of Tampere, Tampere, Finland | | **FinnGen Teams** | **Trajectory** |
| Iida Vähätalo | University of Tampere, Tampere, Finland | | **FinnGen Teams** | **Trajectory** |
| Jyrki Tammerluoto | Institute for Molecular Medicine Finland (FIMM), HiLIFE, University of Helsinki, Helsinl | | **FinnGen Teams** | **Data protection officer** |
| Marco Hautalahti | Finnish Biobank Cooperative - FINBB | | **FinnGen Teams** | **FINBB - Finnish biobank cooperative** |
| Johanna Mäkelä | Finnish Biobank Cooperative - FINBB | | **FinnGen Teams** | **FINBB - Finnish biobank cooperative** |
| Sarah Smith | Finnish Biobank Cooperative - FINBB | | **FinnGen Teams** | **FINBB - Finnish biobank cooperative** |
| Tom Southerington | Finnish Biobank Cooperative - FINBB | | **FinnGen Teams** | **FINBB - Finnish biobank cooperative** |
| Petri Lehto | Finnish Biobank Cooperative - FINBB | | **FinnGen Teams** | **FINBB - Finnish biobank cooperative** |
